# Assessment and Modeling of COVID-19 Outcomes in a Longitudinal Cohort of Hospitalized Adults

**DOI:** 10.1101/2021.07.02.21259665

**Authors:** Lacy M. Simons, Ramon Lorenzo-Redondo, Meg Gibson, Sarah L. Kinch, Jacob P. Vandervaart, Nina L. Reiser, Mesut Eren, Elizabeth Lux, Elizabeth M. McNally, Anat R. Tambur, Douglas E. Vaughan, Kelly E. R. Bachta, Alexis R. Demonbreun, Karla J. F. Satchell, Chad J. Achenbach, Egon A. Ozer, Michael G. Ison, Judd F. Hultquist

**Author notes:** These authors contributed equally to this work (Lacy M. Simons, Ramon Lorenzo-Redondo).

## Abstract

**Background:** While several demographic and clinical correlates of Coronavirus Disease 2019 (COVID-19) outcome have been identified, they remain imprecise tools for clinical management of disease. Furthermore, there are limited data on how these factors are associated with virological and immunological parameters over time.

**Methods and Findings:** Nasopharyngeal swabs and blood samples were longitudinally collected from a cohort of 58 hospitalized adults with COVID-19 in Chicago, Illinois between March 27 and June 9, 2020. Samples were assessed for SARS-CoV-2 viral load, viral genotype, viral diversity, and antibody titer. Demographic and clinical information, including patient blood tests and several composite measures of disease severity, were extracted from electronic health records. All parameters were assessed for association with three patient outcome groups: discharge without intensive care unit (ICU) admission (n = 23), discharge with ICU admission (n = 29), and COVID-19 related death (n = 6). Higher age, male sex, and higher body mass index (BMI) were significantly associated with ICU admission. At hospital admission, higher 4C Mortality scores and lactate dehydrogenase (LDH) levels were likewise associated with ICU admission. Longitudinal trends in Deterioration Index (DI) score, Modified Early Warning Score (MEWS), and serum neutrophil count were also associated with ICU admission, though only the retrospectively calculated median DI score was predictive of death. While viral load and genotype were not significantly associated with outcome in this study, viral load did correlate positively with C-reactive protein levels and negatively with D-dimer, lymphocyte count, and antibody titer. Intra-host viral genetic diversity resulted in changes in viral genotype in some participants over time, though intra-host evolution was not associated with outcome. A stepwise-generated multivariable model including BMI, lymphocyte count at admission, and neutrophil count at admission was sufficient to predict outcome with a 0.82 accuracy rate in this cohort.

**Conclusions:** These studies suggest that COVID-19 disease severity and poor outcomes among hospitalized patients are likely driven by dysfunctional host responses to infection and underlying co-morbid conditions rather than SARS-CoV-2 viral loads. Several parameters, including 4C mortality score, LDH levels, and DI score, were ultimately predictive of participant outcome and warrant further exploration in larger cohort studies for use in clinical management and risk assessment. Finally, the prevalence of intra-host diversity and viral evolution in hospitalized patients suggests a mechanism for population-level change, further emphasizing the need for effective antivirals to suppress viral replication and to avoid the emergence of new variants.

## INTRODUCTION

The emergence of severe acute respiratory syndrome coronavirus 2 (SARS-CoV-2) in late 2019 and its subsequent pandemic spread has ignited a global health crisis. Over 174 million cases of Coronavirus disease 2019 (COVID-19) have been reported as of June 2021, accounting for over 3.7 million deaths worldwide [1]. SARS-CoV-2 infection initiates in the airway epithelia of the upper respiratory tract, usually presenting with symptoms typical of a respiratory illness, including fever, fatigue, cough, shortness of breath, and loss of taste and/or smell [1]. Nevertheless, clinical presentation can range widely from asymptomatic infection to more severe symptoms, including hypoxia and chest pressure [2]. Additional infection and inflammation of the lower respiratory tract can result in life-threatening pneumonia and acute respiratory distress syndrome (ARDS) [3]. Therefore, while most cases are mild, some require clinical intervention in a hospital setting [4].

Understanding risk factors associated with severe disease, hospitalization, and death are crucial for understanding and managing populations most at risk and for improving clinical care of COVID-19 patients. Several studies early in the pandemic identified a number of demographic factors associated with severe COVID-19 including sex (male), age (65 or older), and race/ethnicity (Black, Hispanic, Native American) [5]. Comorbidities and other preexisting conditions - including diabetes, heart disease, asthma, high body mass index (BMI), and immunodeficiency - have likewise been linked with worse clinical outcomes [6–8]. Socioeconomic status, access to healthcare, and exposure risk also have large impacts on risk of transmission and disease [9].

A variety of composite scores have been developed to help predict clinical outcomes and inform medical management and level of care [10]. These scores are typically calculated from several clinical/demographic assessments, including age, BMI, co-morbid conditions, oxygenation status, mental acuity, symptom profile, and laboratory testing. They are designed to aid clinicians in the identification of deteriorating patients with the ultimate goal of improving clinical outcomes. Several of these scores are often used in assessing hospitalized COVID-19 patients, including: the 4C mortality score [11, 12], the Epic Deterioration Index (DI) model [13], and the Modified Early Warning Score (MEWS) [14]. Serial measurement of these scores can be useful for monitoring a patient’s disease progression over time, though they have not been directly compared to one another for their relative clinical utility in assessing COVID-19 outcome.

Besides patient demographics and composite clinical scores, more direct measurements of SARS-CoV-2 infection have similarly been examined as potential biomarkers for disease severity and outcomes. Early reports suggested that viral load in the upper respiratory tract may be a significant predictor of severe disease [15, 16], though this has not been observed in other studies [17]. Likewise, while several new viral lineages have evolved with enhanced transmission or fitness [18–21], few associations have been found between these new genetic variants and disease severity, presentation, or outcomes [22–25]. The host response to infection, on the other hand, has been linked to disease severity in a number of ways. Several individual clinical lab results that monitor the host immune response correlate with COVID-19 disease severity, including lymphopenia, elevated levels of proinflammatory cytokines, and elevated C-reactive protein (CRP) [26, 27]. Individuals with defects in innate immune signaling or who evolve autoantibodies against type I interferons likewise have increased risk for severe disease [28]. Furthermore, serological testing for antibodies against SARS-CoV-2 Spike (S) or Nucleoprotein (N) has suggested that a stronger response to N over S correlates with more severe disease in retrospective analysis [29, 30].

While a number of specific demographic, clinical, virological, and serological measurements have been independently associated with COVID-19 severity and outcomes, the success of these parameters as predictors has been highly variable. It is furthermore unclear how longitudinal assessment of virologic or serologic data might improve composite measures and models of COVID-19 outcome, particularly among hospitalized individuals. Towards this end, we established a biobank of nasopharyngeal swabs and blood samples collected longitudinally from a cohort of hospitalized adults with COVID-19. Serological and virological information from these samples, including viral load, viral genotype, viral diversity, and antibody levels, were analyzed for associations with common clinical metrics of disease severity and participant outcome. Ultimately, these findings will help inform the design and parameters of predictive tools in clinical care and management of COVID-19.

## METHODS

### Specimen collection and processing

After IRB approval, individuals over the age of 18 admitted to Northwestern Memorial Hospital with a positive, PCR-based COVID-19 diagnostic test, who provided informed consent themselves or through an appropriate surrogate, were enrolled in the study. Nasopharyngeal swabs were collected from study participants on the enrollment date and every 4 ± 1 days after enrollment up to 30 days of hospitalization. Swabs were stored in 2-3mL of Primestore MTM (Longhorn Vaccines & Diagnostics, San Antonio, Texas). The Primestore MTM was aliquoted into 2-3 vials of 1mL per tube and stored at -80°C. A total of 238 nasopharyngeal specimens were collected from 58 participants throughout the course of the study. Whole blood was collected from study participants every 8 ± 1 days after enrollment up to 30 days of hospitalization in Vacutainer CPT Mononuclear Cell Preparation tubes containing sodium heparin (Becton Dickinson). Three CPT tubes containing roughly 8 ml of whole blood were collected per participant per time point. A total of 65 whole blood specimens were collected from 34 participants throughout the course of the study; 24 participants either declined consent to blood collection or were unable to provide it at the time of collection. Briefly, sample tubes containing whole blood were centrifuged at 1500-1800 xg in a swing-bucket centrifuge for 30 minutes. Plasma from each of the three collection tubes was removed, pooled, and frozen in 1mL aliquots at -80°C. Peripheral blood mononuclear cells (PBMCs) were removed, washed with 1x PBS containing 0.5% BSA and 2mM EDTA, and frozen in cryopreservation media (1x FBS, 10% DMSO) at -80°C.

### Clinical Data Extraction

Early in the COVID-19 pandemic we obtained IRB approval to create a data mart of all adults diagnosed and treated for COVID-19 across Northwestern Medicine (NM) using the NM Enterprise Data Warehouse (NMEDW) to study the epidemiology, presentation, laboratory/radiographic findings, treatment, co-morbidities, and outcomes of COVD-19. The NMEDW is a joint initiative across Northwestern University Feinberg School of Medicine and Northwestern Memorial Healthcare Corporation to create a single, comprehensive, and integrated repository of all clinical and research data sources to facilitate research, clinical quality, and healthcare operations. The following electronic data elements were extracted and complied once a week from the NMEDW for all adults diagnosed with COVID-19: demographics, health system visits/level of care (i.e., outpatient, ED, hospital, ICU), vital signs, specific laboratory test results, imaging studies, co-morbid conditions/diagnoses (via ICD9/ICD10 coding), pharmacologic therapy (initially via NMEDW pharmacy/medication records then confirmed as needed with electronic chart review), and oxygen delivery/respiratory therapies (initially via NMEDW respiratory therapy records or CPT codes and then confirmed as needed with electronic chart review). From March 1 to June 15, 2020, we also performed electronic chart review and extraction to verify certain NMEDW data (as above) and to collect information on COVID-19 specific symptoms, symptom onset, exposures, and high-risk behaviors. For this study, we linked all available NMEDW and electronic chart review data to this subset of hospitalized participants who provided longitudinal specimens (nasopharyngeal swabs and whole blood) as per the study population and protocol described above. These NMEDW data were utilized to determine demographics, clinical assessments, symptom onset, laboratory measurements, COVID-19 disease severity, and hospital outcomes (ICU care and death) for the study analyses.

### Viral Load Determination

Viral RNA was extracted from nasopharyngeal specimens utilizing the QIAamp Viral RNA Minikit (Qiagen). Laboratory testing for SARS-CoV-2 presence was performed by quantitative reverse transcription and PCR (qRT-PCR) with the CDC 2019-nCoV RT-PCR Diagnostic Panel utilizing N1 and RNase P probes as previously described [31]. Specimens with undetectable RNase P levels were considered to be of insufficient quality and were excluded from future studies. Ct values from the N1 probes were used in all subsequent analyses with N1 Ct values less than or equal to 35 considered positive.

### cDNA Synthesis and Viral Genome Amplification

cDNA synthesis was performed with SuperScript IV First Strand Synthesis Kit (Thermo) using random hexamer primers according to manufacturer’s specifications. Direct amplification of the viral genome cDNA was performed in multiplexed PCR reactions to generate ∼400 base pair amplicons tiled across the genome. The multiplex primer set, comprised of two non-overlapping primer pools, was created using Primal Scheme and provided by the Artic Network (version 3 release). PCR amplification was carried out using Q5 Hot Start HF Taq Polymerase (NEB) with 5 µl of cDNA in a 25 µl reaction volume. A two-step PCR program was used with an initial step of 98°C for 30 seconds, then 35 cycles of 98°C for 15 seconds followed by five minutes at 65°C. Separate reactions were carried out for each primer pool and validated by agarose gel electrophoresis.

### Sequencing Library Preparation and Illumina Sequencing

The sequencing library approach was adapted from previously published methods [32]. Briefly, amplicons from both primer pools were combined and purified with a 1x volume of AmpureXP beads (Beckman Coulter). A total of 75 ng of DNA was treated with KAPA HyperPrep End Prep Enzyme mix (KAPA). Up to 96 specimen libraries were barcoded using NEXTflex barcodes and KAPA HyperPrep DNA Ligase (KAPA) for simultaneous sequencing. Uniquely barcoded samples were pooled and purified with a 0.8x volume of AmpureXP beads. Library amplification was performed using KAPA HiFi HotStart with KAPA Library Amp Primers. Amplicons were purified with a 0.8x volume of AmpureXP beads and normalized to 5 nM and pooled. The pooled library was denatured and loaded onto a MiSeq v2 500 cycle flow cell (Illumina). Viral genome consensus sequences were determined from sequencing reads as previously described [33]. Sequencing reads were aligned to the reference SARS-CoV-2 genome sequence MN908947.3 using bwa version 0.7.15. Barcode sequences were trimmed from aligned reads and consensus sequence determined using iVar v1.2.2 [33] with a minimum alignment depth of 10 reads and minimum base quality of 20, and a consensus frequency threshold of 0 (i.e. majority base as the consensus). Consensus sequences with ≥ 10% missing bases were discarded.

### SARS-CoV-2 Nucleocapsid Protein Expression and Purification for ELISA

For expression of the SARS-CoV-2 Nucleocapsid (N-protein) N-terminal domain (NTD, amino acids 47-173) and C-terminal domain (CTD, amino acids 247-364), the corresponding DNA sequence from the Wuhan-Hu-1 nucleocapsid open reading frame [34] was codon optimized for expression in *Escherichia coli* (*E. coli)* using Genscript Gensmart and Bio Basic software, respectively. The NTD sequence was synthesized (Twist Bioscience) and cloned into pMCSG53 [35], yielding pMCSG53-N-NTD (available through BEI Resources, NR-52428). The CTD sequence was synthesized (Bio Basic) and cloned into pET11a (Novagen), yielding pET11a-N-CTD (available through BEI Resources, NR-52434). These expression plasmids were transformed into BL21(DE3) *E.coli* cells [36]. For expression of full-length SARS-CoV-2 N-protein (N-FL, amino acids 1-412), pET-28a(+) containing the entire Wuhan-Hu-1 Nucleocapsid open reading frame was obtained from BEI Resources (NIAID, NIH, Catalog # NR-53507) and transformed into *E. coli* Rosetta-1 cells (Novagen). *E.coli* expressing each of the three plasmids were independently grown overnight under appropriate selection and protein expression was induced with isopropyl-d-thiogalactopyranoside (IPTG). For all three proteins, *E. coli* cells were harvested by centrifugation at 6,000 RCF for 10 minutes. Cell pellets were resuspended in lysis buffer (500 mM NaCl, 10mM Tris pH 7.5, and 5% glycerol) and sonicated (5 seconds on, 10 seconds off, 10% amplitude for 20 minutes) using a Qsonica Q500 Sonicator. The sonicated lysate was centrifuged at 18,000 RCF for 40 minutes and the supernatant was purified on an ÄKTA Pure fast protein liquid chromatography (FPLC) system using a 5 mL GE Healthcare HisTrapFF nickel (Ni^2+^) affinity column. The column was first equilibrated with loading buffer (0.5 M NaCl, 10 mM tris(hydroxymethyl)aminomethane (Tris) pH 7.5, 1 mM tris2-carboxyethylphosphine (TCEP)). After sample application, the column was washed with loading buffer followed by a high salt buffer (1 M NaCl, 10 mM TRIS pH 7.5, 25 mM imidazole, 0.1 mM TCEP) and purified proteins were eluted with elution buffer (0.5 M NaCl, 10 mM Tris pH 7.5, 0.5 M imidazole, 0.1 mM TCEP). Eluted protein was dialyzed overnight in loading buffer. Purified protein was aliquoted and stored in 50% glycerol at -80°C.

### ELISA for SARS-CoV-2 NTD, CTD and N-FL proteins

Human serum samples were centrifuged at 15,000 RCF for 10 minutes. The top layer was harvested and heat inactivated at 55°C for 30 minutes. Following heat inactivation, the serum was centrifuged at 21,000 RCF for 30 minutes at 4°C and stored at -20°C. Briefly, 96-well microtiter plates (Nunc-Immuno™MicroWell™96 well solid plates, Fisher) were coated with 100 µL of 5 µg/mL target protein in 1x Phosphate Buffered Saline (PBS) and incubated at 4°C for 48 hours. Coated plates were washed three times with 250 µL of wash buffer (1x PBS with 0.5% Tween-20) using a Thermo Fisher Wellwash™ Versa microplate washer. Plates were then blocked for four hours with 200 µL of blocking buffer (1x PBS, 0.5% Tween-20, 2% Bovine Serum Albumin (Millipore Sigma)). Human serum was diluted 1:1000 in blocking buffer, and 100 µL was added to each well and incubated for one hour at room temperature. Following another three washes, plates were incubated for one hour with 100 µL/well of 1 µg/mL horseradish peroxidase conjugated goat anti-human IgG antibody, F(ab′)2 (Chemicon) in blocking buffer prior to development with 3,3’,5,5’-Tetramethylbenzidine (TMB) solution (Fisher). Development was quenched with 100 µl of 0.2 M sulfuric acid and absorbance was measured at 450 nm (Molecular Devices spectra max m3spec using SoftMax Pro v6.5.1 software). Each serum sample was analyzed in triplicate (n=3) and ELISAs were replicated for each protein (NTD, CTD, N-FL) on two separate days, yielding a total of six data points for each serum sample for each of the three proteins assayed.

### Anti-Spike Antibody Quantification by ELISA

Anti-Spike IgG concentration in serum samples was determined utilizing an ELISA assay as described [37, 38]. In brief, 96-well plates (ThermoFisher Scientific #3855) were coated with 2 µg/mL RBD antigen overnight. Plates were washed and blocked for 2 hrs. Diluted serum was transferred to the coated 96-well plate and incubated for 60 mins. SIGMA*FAST™* OPD (Sigma-Aldrich #P9187) was prepared and 100µl added to each well for 10 mins. 3M HCL was used as a stop solution. Absorbance (optical density, OD) was read at 490 nm (BioTek Synergy H1). All samples dilutions were run in duplicate and reported as an average. A four-parameter logistic regression of the multi-concentration standard curve was obtained using CR3022, a recombinant human anti-SARS-CoV-RBD IgG antibody, with a known affinity to the RBD of SARS-CoV-2 (CR3022 antibody, Creative Biolabs #MRO-1214LC). The standard curve was used to calculate the anti-RBD IgG concentration (µg/ml) in serum samples. A value >0.39µg/ml CR3022 was considered positive as described [38].

### Anti-HLA Antibody Quantification

The presence of HLA Class I and Class II antibodies was measured using the FlowPRA™ Screening assay (One Lambda, A Thermo Fisher Scientific Brand), following manufacturer’s recommendation. Each of the FlowPRA Class I and Class II assays consist of a pool of 30 different bead preparations, representing all common antigens as well as many rare HLA alleles. Participant serum was centrifuged to remove any aggregates and incubated with Class I and II beads to bind. Class I and Class II antibodies were screened simultaneously using flow cytometry (Cytoflex, Beckman Coulter). Histograms of participants’ sera were compared to the negative control and the positive population was gated to determine percent positivity.

### PAI-1 Quantification by ELISA

To measure the plasma levels of human plasminogen activator inhibitor type-1 (PAI-1), we performed a sandwich ELISA method that detects free, latent, and complexed forms of PAI-1 using a Human PAI-1 total antigen assay ELISA kit (Molecular Innovations, Catalog #: HPAIKT-TOT). Following the manufacturer’s recommended protocol, plasma samples were diluted five to ten times in Tris-Buffered Saline (TBS) containing 3% BSA (w/v) before added into the microtiter plate wells containing the PAI-1 capture antibodies. After washing away the unbound fraction of plasma samples, the amount of PAI-1 captured in the wells was quantified after sequential binding, washing, incubation of primary antibody against PAI-1, incubation of the secondary antibody-HRP conjugate and development utilizing a TMB substrate. The optical density was measured at 450 nm on a plate reader.

### Phylogenetic Analysis

Consensus sequences for all longitudinal samples from each participant were aligned using MAFFT v7.453 software and manually edited using MEGA v6.06. A Maximum Likelihood (ML) phylogeny with all consensus sequences were inferred with IQ-Tree v2.0.5 using its ModelFinder function before each analysis to estimate the nucleotide substitution model best-fitted for each dataset by means of Bayesian information criterion (BIC). We assessed the tree topology for each phylogeny both with the Shimodaira–Hasegawa approximate likelihood-ratio test (SH-aLRT) and with ultrafast bootstrap (UFboot) with 1000 replicates each. SARS-CoV-2 clades were assessed using Nextclade (clades.nextstrain.org) and Pango lineages were assigned to the consensus sequences using pangolin software [39]. Nucleotide changes along the tree branches were identified by ML ancestral reconstruction using TreeTime v0.7.6.

### Viral Diversity Analysis

All trimmed sequencing reads obtained were also used to reconstruct the SARS-CoV2 haplotypes for all SARS-CoV-2 genes following genome sequence MN908947.3 location in every sample with successful viral amplification. We used QuasiRecomb [40] to perform a probabilistic inference of the viral haplotypes per gene present in each viral population. After haplotype reconstruction, we discarded sequences with less than 1% frequency of the reads to avoid including sequencing errors and we calculated the pairwise genetic distance between every haplotype and the most predominant haplotype using DistanceCalculator in biopython 1.74. Following genetic distance calculation, values were weighted by haplotype frequency and a weighted average diversity value was obtained by the sum of all the values obtained per gene for every sample. To obtain the number of nonsynonymous substitutions per non-synonymous site (dN) and the number of synonymous substitutions per synonymous site (dS), we followed a similar approach using CodonSeq and cal_dn_ds with the NG86 method from biopython 1.74. These values were weighted by the frequency of the haplotype and the difference dN-dS was calculated. The low levels of intra-host diversity in the collected SARS-CoV-2 isolates resulted in a high number of ‘0’ values that precluded the use of dN/dS ratios in this study, so dN-dS values were calculated instead. All calculations were performed using in-house scripts in python 3.8 (available upon request).

### Statistics and Modeling

All statistical analyses and modeling were performed using R version 4.0.3. All simple correlations were performed using Spearman’s rank correlation. Pairwise group comparisons were performed using Wilcoxon rank sum test followed by the Benjamini-Hochberg procedure to control the False Discovery Rate (FDR) for multiple comparisons. A FDR<0.05 was used as statistical significance cut-off. Initial modeling of outcome as a function of demographic predictor was performed by fitting a multinomial log-linear model using nnet package followed by chi-squared tests to examine the contributions of the individual factors. We included all demographic factors as well as the comorbidity score in the fitted model to examine which of these factors significantly contributed to the observed outcome. To cluster participants according to their N antibody levels, we used Principal Component Analysis (PCA) where we included all replicate values obtained for each of the three antibody types analyzed. The clustering of each study participant was subsequently obtained by agglomerative hierarchical clustering on the PCA results. After building and initial hierarchical tree, the sum of within-cluster inertia was calculated for each partition. The selected partition was the one with the higher relative loss of inertia. Both PCA and agglomerative hierarchical clustering were performed using FactoMineR package and factoextra was used for visualization of the clustering results. Multinomial logistic models were performed only in datasets with complete cases using nnet package and stepwise model selection was performed by Akaike Information Criterion (AIC) both using forward inclusion and backward elimination of variables using MASS package.

## RESULTS

### Demographic and clinical characteristics of the cohort

A total of 63 participants admitted to Northwestern Memorial Hospital with a positive COVID-19 diagnostic test were enrolled in the study between March 27, 2020 and June 9, 2020 during the first wave of the pandemic in Chicago. Of these participants, 58 provided at least one nasopharyngeal swab and 32 provided at least one blood specimen (**Figure 1A**). Four participants ultimately had no samples collected while one participant declined nasopharyngeal swab collection; these participants were excluded from further analysis. The cohort skewed slightly male (36 male, 22 female), consistent with broader COVID-19 hospitalization trends [41] (**Table 1** and **Figure 1B**). Age of the participants was roughly equivalent by sex with an average age of 63.6 years (S.D. = 15.2) among males and 62 years (S.D. = 18.2) among females (**Table 1** and **Figure 1B**). Additional demographic and clinical data are summarized in **Table 1**.

**Figure 1.**
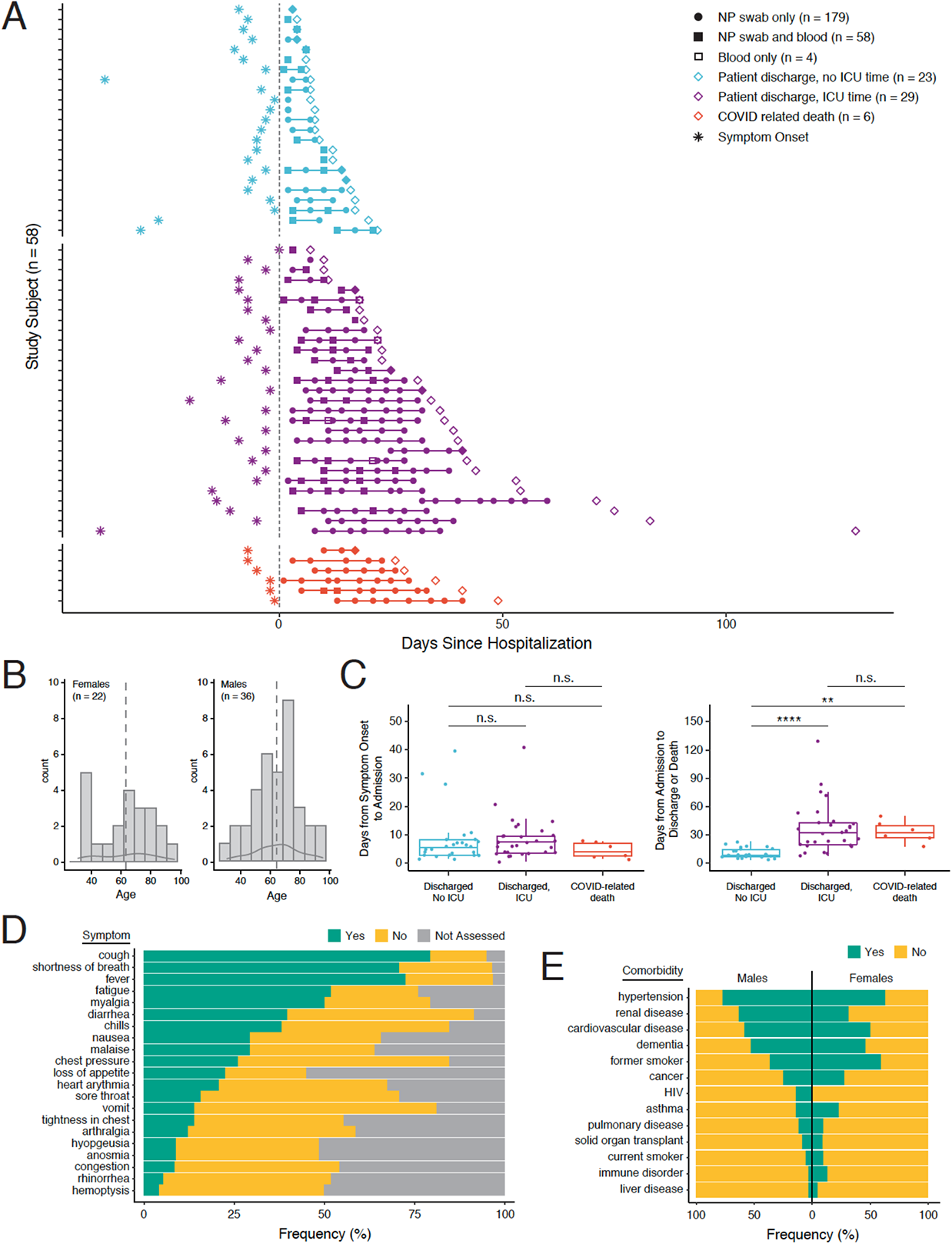
Cohort Description and Longitudinal Sampling Strategy. **A)** A graphical representation of samples taken per study participant along the clinical time course of inpatient treatment for COVID-19 (n = 58 total participants). Participants are grouped by outcome: discharge with no ICU care required (blue, n = 23), discharge with come ICU care required (purple, n = 29), and COVID-related death (red, n = 6). Sample collection is bracketed by symptom onset (asterisk) and hospital discharge or death (diamond) with the dotted line representing the time of hospital admission. Nasopharyngeal swabs (closed circles), blood (open squares), or both (closed squares) were collected from each study participant as indicated. **B)** Age distribution of study participants by sex with median age depicted by the dotted line. **C)** Box plots comparing the time between symptom onset and hospital admission and the time between hospital admission and discharge or death by outcome (Wilcoxon rank sum test with Benjamini-Hochberg procedure to control False Discovery Rate (FDR) for multiple comparisons, n.s. = FDR >0.05, ** = FDR <0.005, **** = FDR <0.00005). **D)** Frequency of reported symptoms among study participants during hospitalization ranked by most frequently to least frequently reported (green = present, yellow = not present, gray = not assessed). **E)** Frequency of reported symptoms among study participants by sex (green = present, yellow = not present).

**TABLE 1.** Summary of demographics and comorbidities of study participants.

| Variable | Descriptor | Female | Male |
| --- | --- | --- | --- |
| Total N (%) | - | 22 (37.9) | 36 (62.1) |
| Age | Mean (SD) | 62 (18.2) | 63.6 (15.2) |
| Blood type | n/a | 7 (31.8) | 10 (27.8) |
|  | A Negative | 1 (4.5) | 1 (2.8) |
|  | A Positive | 5 (22.7) | 4 (11.1) |
|  | AB Positive | 1 (4.5) | 2 (5.6) |
|  | B Positive | 1 (4.5) | 2 (5.6) |
|  | O Negative | 0 (0.0) | 2 (5.6) |
|  | O Positive | 7 (31.8) | 15 (41.7) |
| Race | Asian | 0 (0.0) | 1 (2.8) |
|  | Black or African American | 10 (45.5) | 15 (41.7) |
|  | Declined | 1 (4.5) | 3 (8.3) |
|  | Other | 3 (13.6) | 2 (5.6) |
|  | White | 8 (36.4) | 15 (41.7) |
| Ethnicity | Declined | 0 (0.0) | 2 (5.6) |
|  | Hispanic or Latino | 4 (18.2) | 7 (19.4) |
|  | Not Hispanic or Latino | 18 (81.8) | 27 (75.0) |
| Body Mass Index (BMI) | Mean (SD) | 38.6 (13.0) | 31.3 (7.4) |
| Asthma | No | 17 (77.3) | 31 (86.1) |
|  | Yes | 5 (22.7) | 5 (13.9) |
| COPD | No | 20 (90.9) | 32 (88.9) |
|  | Yes | 2 (9.1) | 4 (11.1) |
| Cancer | No | 16 (72.7) | 27 (75.0) |
|  | Yes | 6 (27.3) | 9 (25.0) |
| Cardiovascular Disease | No | 11 (50.0) | 15 (41.7) |
|  | Yes | 11 (50.0) | 21 (58.3) |
| Chronic Liver Disease | No | 21 (95.5) | 35 (97.2) |
|  | Yes | 1 (4.5) | 1 (2.8) |
| Diabetes | No | 12 (54.5) | 17 (47.2) |
|  | Yes | 10 (45.5) | 19 (52.8) |
| HIV Diagnosis | No | 22 (100.0) | 31 (86.1) |
|  | Yes | 0 (0.0) | 5 (13.9) |
| Hypertension | No | 8 (36.4) | 8 (22.2) |
|  | Yes | 14 (63.6) | 28 (77.8) |
| Immune Disorder | No | 19 (86.4) | 35 (97.2) |
|  | Yes | 3 (13.6) | 1 (2.8) |
| Renal Disease | No | 15 (68.2) | 13 (36.1) |
|  | Yes | 7 (31.8) | 23 (63.9) |
| Solid Organ Transplant | No | 20 (90.9) | 33 (91.7) |
|  | Yes | 2 (9.1) | 3 (8.3) |
| Former Smoker | No | 9 (40.9) | 23 (63.9) |
|  | Yes | 13 (59.1) | 13 (36.1) |
| Current Smoker | No | 20 (90.9) | 34 (94.4) |
|  | Yes | 2 (9.1) | 2 (5.6) |

Note: Data are represented as Mean (SD) or N (%) for continuous or categorical variables respectively.
COPD = Chronic Obstructive Pulmonary Disorder
HIV = Human Immunodeficiency Virus

Based on overall clinical course, we utilized NMEDW data (described above) to categorize participants into three hospital outcome groups: discharged without being admitted to the Intensive Care Unit (ICU) (n = 23); discharged after admission to the ICU for some period during their hospital stay (n = 29); and death due to COVID-19 related illness (n = 6). The time between symptom onset and hospital admission was not significantly different between different outcome groups (**Figure 1C**), though total length of stay was significantly higher for participants admitted to the ICU or who died (**Figure 1C**). The most frequent symptoms prior to admission were cough, shortness of breath, fever, fatigue and myalgia, each of which was reported by over 50% of participants (**Figure 1D**). Most participants had at least one underlying co-morbid condition with hypertension, renal disease, and cardiovascular disease being the most frequent (**Figure 1E**). Overall, more males than females were admitted to the ICU (two-thirds of males and one-half of females, **Table 2**), consistent with reports from other hospital systems [42].

**TABLE 2.**
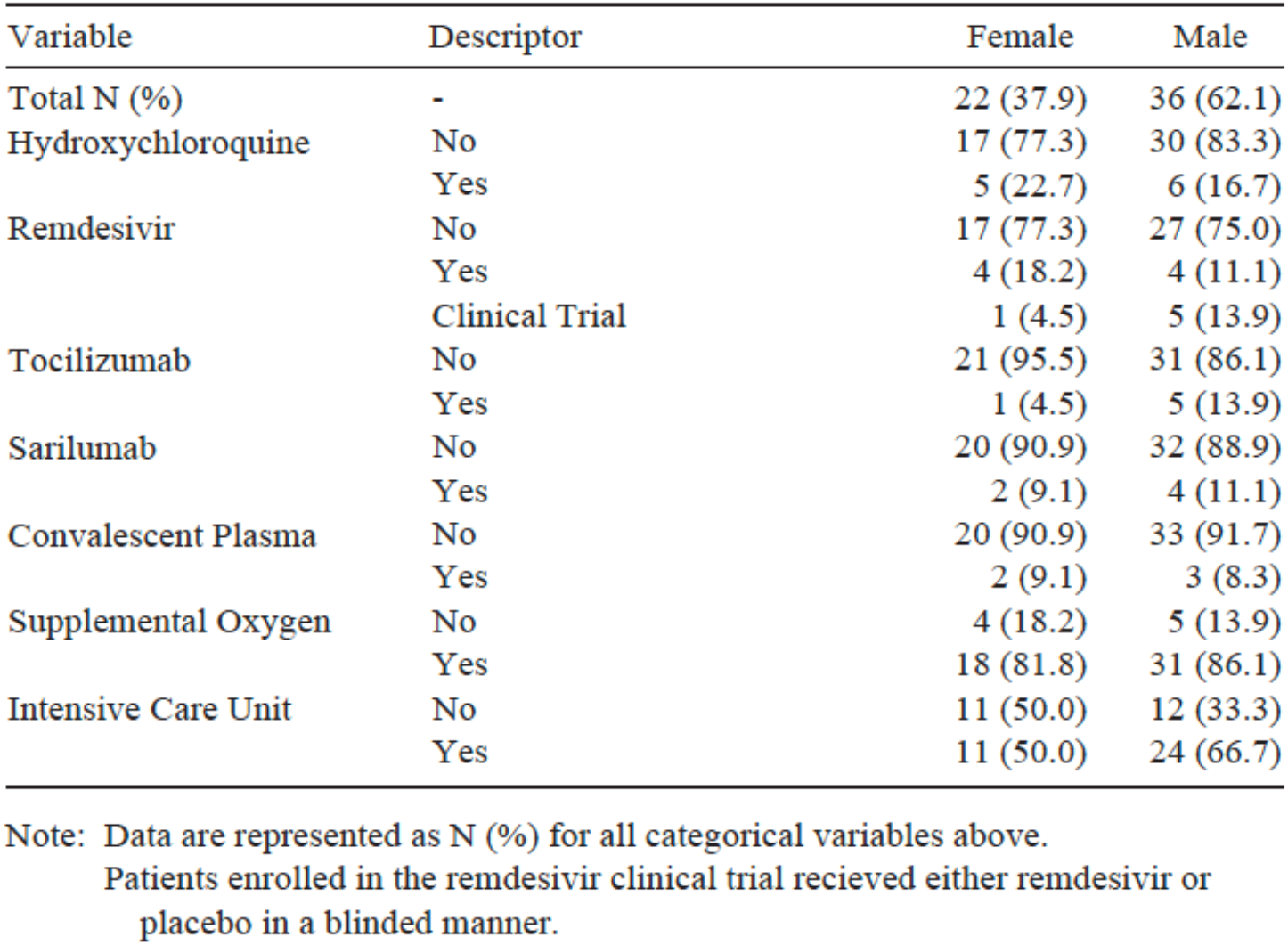
Summary of treatments administered to study participants during hospitalization.

| Variable | Descriptor | Female | Male |
| --- | --- | --- | --- |
| Total N (%) | - | 22 (37.9) | 36 (62.1) |
| Hydroxychloroquine | No | 17 (77.3) | 30 (83.3) |
|  | Yes | 5 (22.7) | 6 (16.7) |
| Remdesivir | No | 17 (77.3) | 27 (75.0) |
|  | Yes | 4 (18.2) | 4 (11.1) |
|  | Clinical Trial | 1 (4.5) | 5 (13.9) |
| Tocilizumab | No | 21 (95.5) | 31 (86.1) |
|  | Yes | 1 (4.5) | 5 (13.9) |
| Sarilumab | No | 20 (90.9) | 32 (88.9) |
|  | Yes | 2 (9.1) | 4 (11.1) |
| Convalescent Plasma | No | 20 (90.9) | 33 (91.7) |
|  | Yes | 2 (9.1) | 3 (8.3) |
| Supplemental Oxygen | No | 4 (18.2) | 5 (13.9) |
|  | Yes | 18 (81.8) | 31 (86.1) |
| Intensive Care Unit | No | 11 (50.0) | 12 (33.3) |
|  | Yes | 11 (50.0) | 24 (66.7) |

Note: Data are represented as N (%) for all categorical variables above.
Patients enrolled in the remdesivir clinical trial recieved either remdesivir or placebo in a blinded manner.

Between March and June of 2020, the standard of care and available treatments for COVID-19 patients were changing rapidly, with some participants of this study concurrently enrolling in double-blinded clinical trials [43, 44]. For participants who were in clinical trials, treatment arm was not available to the study team to incorporate into this analysis. While a majority of participants of both sexes received supplemental oxygen (81.8% of females and 86.1% of males), no other treatment was prescribed to more than 25% of study participants (**Table 2**).

### Demographic associations with COVID-19 outcome

Previous studies have identified several demographic characteristics that are associated with increased risk of severe COVID-19 and worse outcome, including sex (male), age (>65), race (Black or African American), ethnicity (Hispanic or Latino), body mass index (BMI >25), and the presence of underlying co-morbidities (>1) [45, 46]. To assess for correlations between these demographic parameters and outcome in our cohort, we constructed a logistic regression model with categorical outcome (non-ICU, ICU, or death) as our dependent variable and sex, age, race, ethnicity, BMI, and total number of reported co-morbidities as our independent variables (**Figure 2A**). Sex (p-value = 0.0038; **Figure 2B**), age (p-value = 0.0004; **Figure 2C**), and BMI (p-value = 0.0009; **Figure 2D**) were significantly associated with ICU admission, while race, ethnicity, and co-morbidities were not correlated with outcome group in this study.

**FIGURE 2.**
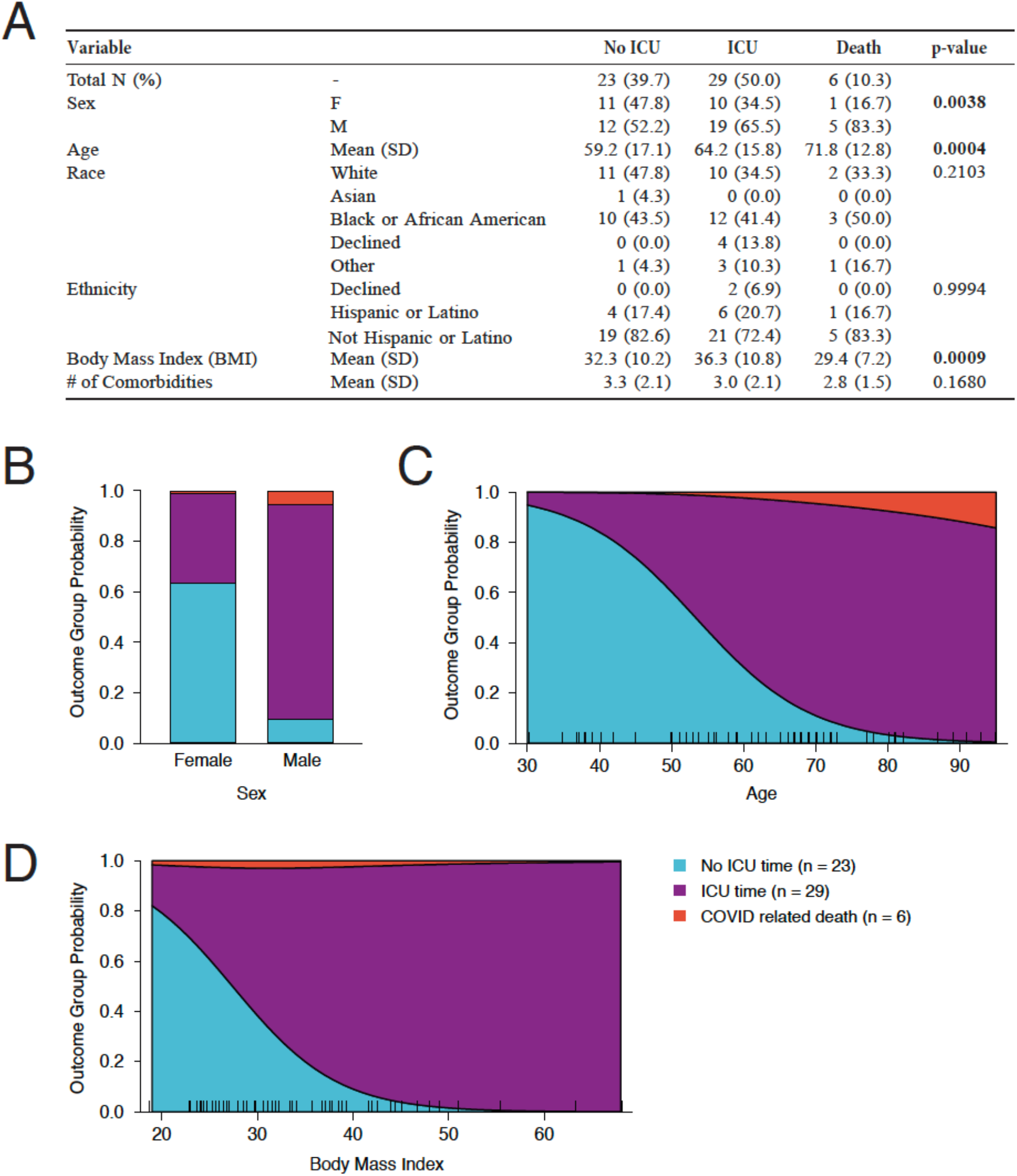
Multinomial Logistic Regression Model of Participant Outcome by Demographic and Comorbidities. **A)** Distribution and statistical significance of study participant sex, age, race, ethnicity, body mass index (BMI), and comorbidity index towards predicting outcome as defined by never requiring ICU care, requiring some ICU care, or COVID-related death. Categorical variables are presented as count (percent) while continuous variables are represented as mean (standard deviation). Significant contributors to the logistic model (p-value < 0.05) are bolded. **B)** Probability of each outcome determined by sex in the study cohort per the logistic model. **C)** Probability of each outcome determined by age in the study cohort per the logistic model. The age of each study participant is indicated by a tick mark along the x-axis. **D)** Probability of each outcome determined by BMI in the study cohort per the logistic model. The BMI of each study participant is indicated by a tick mark along the x-axis.

### Associations between composite clinical scores and COVID-19 outcome

Three composite measures of disease severity were calculated for each participant at admission and/or longitudinally over their entire hospital stay: the Epic Deterioration Index (DI) score [13], Modified Early Warning score (MEWS) [14] and the 4C Mortality score (4C) [11, 12]. The DI score is a proprietary severity score calculated daily by Epic software developed with the goal of identifying patients who are deteriorating and require more intensive intervention and/or care [47]. It ranges from 0 to 100 with higher scores reflective of greater risk of adverse outcomes. Over the course of their hospital stay, the non-ICU group had consistently lower DI scores than the ICU and death groups, with most participants peaking at or below a score of 50 (**Figure 3A**). Although individuals who required ICU care tended to have higher DI scores at hospital admission (No ICU, median DI = 30.30; ICU, median DI = 39.55; COVID-related death, median DI = 45.75; **Supplemental Figure 1A**), they were not significantly different by pairwise comparison. Over their first 10 days of hospitalization, however, participants requiring ICU care exhibited steep increases in DI score, suggestive of rapid deterioration (**Figure 3B**). After the first 10 days, participants who ultimately died saw slow, but steady increases in DI score while participants who recovered from the ICU saw a lowering of the DI score after a stochastic interval (**Figure 3A** and **3B**). Reflective of these trends, the median DI score over the participant’s entire hospital stay was significantly associated with each outcome group (**Figure 3C**). The minimum and maximum recorded DI score over the participant’s hospital stay was likewise significantly different between non-ICU and ICU participants, but not significantly different for ICU participants who recovered versus those who died (**Supplemental Figure 1A**).

**FIGURE 3.**
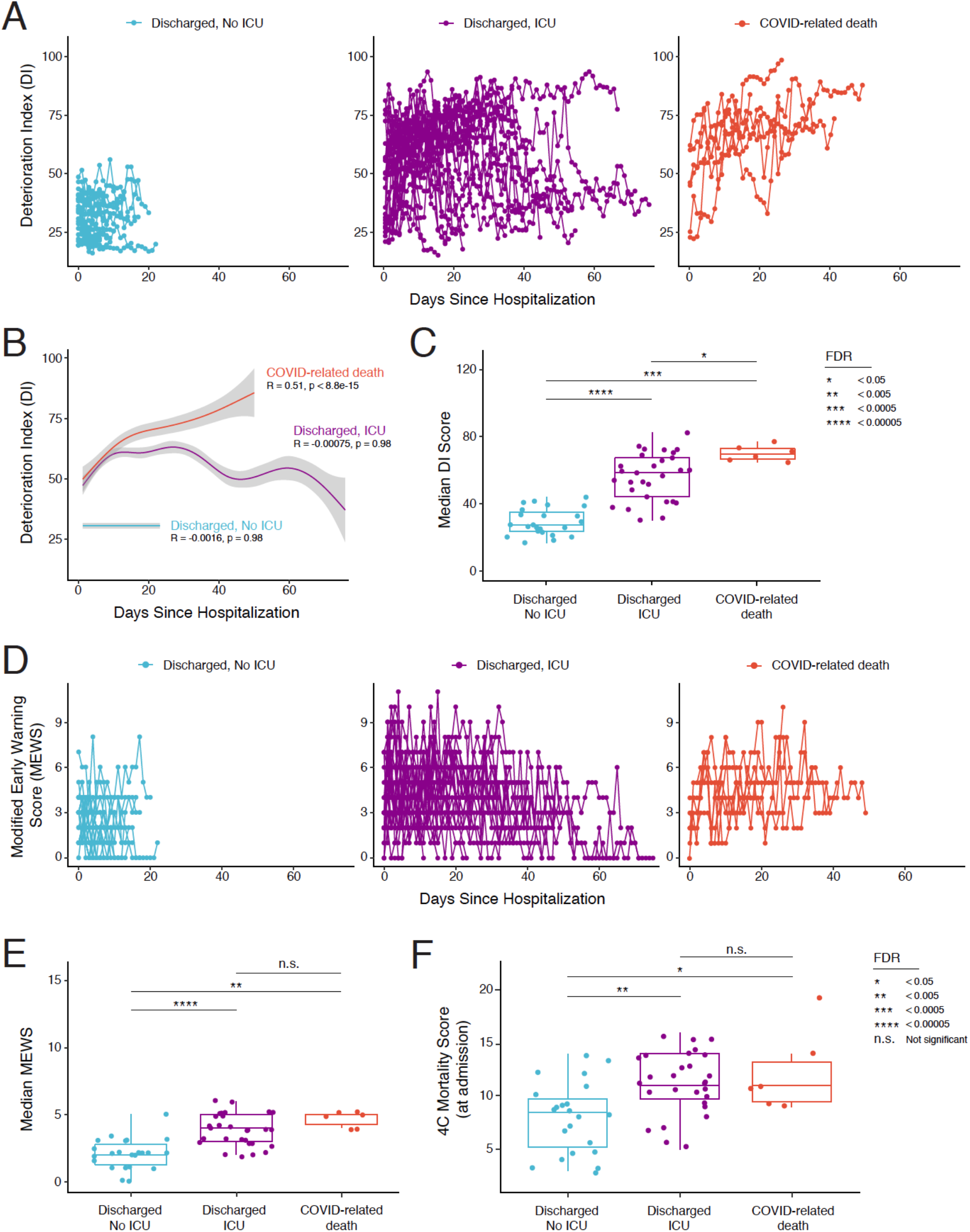
Analysis of correlations between clinical measures of disease severity and participant outcome. **A)** Plot of the Deterioration Index (DI) score for each study participant over the course of hospitalization, separated by participant outcome. **B)** Generalized additive model fit to the average DI score of hospitalized study participants over time separated by outcome. The correlation between average DI score and time during hospitalization is provided (R) alongside the p-value for each outcome group. **C)** Box plot comparing the median reported DI score for each participant grouped by outcome. **D)** Plot of the Modified Early Warning Score (MEWS) for each study participant over the course of hospitalization, separated by participant outcome. **E)** Box plot comparing the median reported MEWS for each participant grouped by outcome. **F)** Box plot comparing the 4C Mortality score measured at admission for each participant grouped by outcome. Significance between groups in all box plots was tested by Wilcoxon rank sum test with Benjamini-Hochberg procedure to control FDR for multiple comparisons; n.s. = FDR >0.05, * = FDR <0.05, ** = FDR <0.005, *** = FDR <0.0005, **** = FDR <0.00005.

The Modified Early Warning score (MEWS) is a clinical severity score that compiles information on systolic blood pressure, heart rate, respiratory rate, temperature, and Alert Voice Pain Unresponsive (AVPU) Score to predict patient deterioration [48]. MEWS ranges from 0 to 14 with higher scores indicative of risk for more adverse outcomes. Over the course of their hospital stay, the non-ICU group tended to have lower MEWS scores than the ICU and death groups, though this difference was less pronounced than with the DI scores (**Figure 3D**). Indeed, while the median and maximum MEWS values reported over a participant’s hospital stay were significantly different between individuals that required ICU care and those that did not (**Figure 3E**, **Supplemental Figure 1B**), there were no significant differences in MEWS values between outcome groups at admission and no significant differences were detected between the ICU and death groups (**Figure 3E**, **Supplemental Figure 1B**).

Finally, the 4C Mortality score was recently developed by the International Severe Acute Respiratory and Emerging Infection Consortium (ISARIC) [12] specifically for risk stratification of COVID-19 patients upon hospital admission. It includes a number of both demographic (sex, age, number of co-morbidities) and clinical (respiratory rate, peripheral oxygen saturation, urea levels, CRP, Glasgow Coma Scale) inputs that are used to generate a score between 0 and 21 with higher scores associated with higher risk of mortality [11]. In our cohort, we found that 4C mortality scores at admission were significantly higher for both the ICU and death outcome groups compared to the non-ICU group, though the 4C Mortality scores between the ICU and death groups were not significantly different (**Figure 3F**).

### Associations between clinical measures and COVID-19 outcome

A number of individual clinical measures and laboratory test results were collected from participants both at the time of admission as well as periodically over the duration of their hospital stay. These include blood cell counts (total white blood cells, lymphocytes, neutrophils, etc.), blood protein levels [C-reactive Protein (CRP), lactate dehydrogenase (LDH), D-dimer, ferritin, procalcitonin, etc.], and measures of oxygenation [fraction of inspired oxygen (FiO_2_) and oxygen saturation (SpO_2_)]. Non-ICU participants had a significantly lower maximum FiO_2_ (28.1+/-8.5%) and a higher minimum SpO_2_ (92.5+/-3.1%) at admission compared to participants who required ICU care (59.4+/-32.4% and 84.2 +/-12.7%, respectively) or who died (69.7+/-36.2% and 83.8+/-11.0%, respectively) (**Figure 4A**). Furthermore, blood levels of LDH, a marker of tissue damage, were significantly higher in ICU participants (455.4+/-220.7 Units/L) compared to non-ICU participants (287.6+/-95.7 Units/L) at admission (**Figure 4A**, **Figure 4B**). No other laboratory test result or clinical measure taken at hospital admission was significantly associated with any outcome group.

**FIGURE 4.**
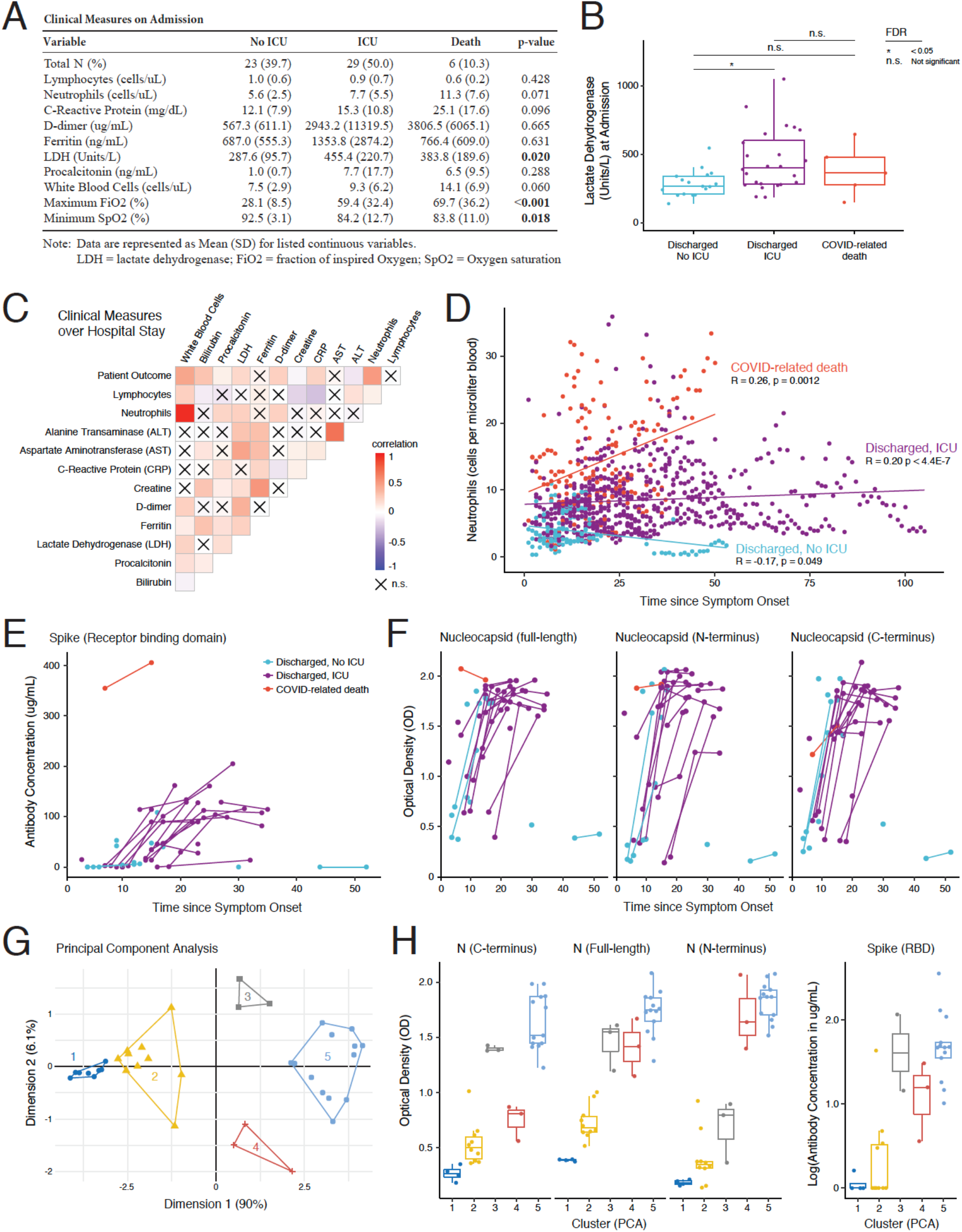
Analysis of correlations between clinical laboratory tests, serology, and participant outcome. **A)** Table summarizing the average value of each variable measured in routine clinical blood work taken at the time of hospital admission by outcome group. All continuous variables are reported as mean (standard deviation). Chi-square test of independence was used to assess differences among the outcome groups (p-values < 0.05 in bold). **B)** Box plot of lactate dehydrogenase (LDH) activity in the blood at admission by outcome group (Wilcoxon rank sum test with Benjamini-Hochberg procedure to control FDR for multiple comparisons, n.s. = FDR >0.05, * = FDR <0.05). **C)** Heat map of correlation among longitudinally reported blood work variables over the entire course of hospitalization alongside outcome group. Value of the correlation coefficient is colored blue to red from more negatively to more positively correlated, respectively. Lack of significant correlation using Spearman correlation (p-value >0.05) is indicated by an X. **D)** Scatterplot of neutrophil count in the blood (cells per microliter) by time since symptom onset. Dots are colored by outcome group (no ICU care in blue, some ICU care in purple, COVID-related death in red) with the linear regression fit shown as a solid line of the same color (Spearman correlation coefficient and p-value shown). **E)** Line graph of the concentration of IgG antibody (micrograms per milliliter) recognizing the receptor binding domain (RBD) of Spike as determined by ELISA per study participant relative to the time since symptom onset. Lines are colored by outcome group. **F)** Line graph of the Optical Density (OD) value reflective of the concentration of antibody recognizing the full-length (left), N-terminus (center), or C-terminus (right) of the Nucleocapsid (N) protein as determined by ELISA per study participant. Values are presented relative to time since symptom onset and lines are colored by outcome group. **G)** Principal component analysis (PCA) plot depicting study participant grouping by N antibody levels obtained by agglomerative hierarchical clustering. **H)** Box plots comparing the OD value reflective of the concentration of antibody recognizing the C-terminus (left), full-length (center), or N-terminus (right) of the N protein as determined by ELISA per PCA grouping. The box plot on the far right depicts the concentration of IgG antibody recognizing Spike RBD in each specimen by PCA grouping.

While LDH was the only clinical blood test result to show a significant association with participant outcome at admission, several factors were associated with outcome when considered over the entire hospital stay (**Figure 4C**). Most notably, increased neutrophil count over the course of hospitalization was significantly associated with worse outcomes. While non-ICU participants had relatively low neutrophil counts that slightly decreased over the course of hospitalization, ICU participants had higher median levels that persisted throughout their stay (**Figure 4D**). Among participants who died, we observed increasing neutrophil counts over the course of hospitalization (**Figure 4D**), consistent with prior reports [49, 50].

While these laboratory tests used in routine clinical care capture some cellular and molecular aspects of the immune response, they fail to capture the potential impact of the humoral response to infection. Both the ratio of anti-Spike to anti-Nucleocapsid antibodies and the increased presence of antibodies to the carboxy (C) terminus of Nucleocapsid have been previously suggested to correlate with COVID-19 outcome [51, 52]. To assess these patterns in our cohort, plasma collected from a subset of participants (**Figure 1A**) was analyzed for the presence of anti-Spike IgG (Receptor binding domain, **Figure 4E**) and anti-Nucleocapsid IgG (N-terminal, C-terminal, and full-length protein, **Figure 4F**) by enzyme linked immunosorbent assays (ELISAs). As expected, most participants developed an antibody response to both the SARS-CoV-2 Spike and Nucleocapsid protein within 10-20 days of symptom onset (**Figure 4E, Figure 4F**). Overall, the response to each antigen was highly correlated across participants (**Supplemental Figure 2D**). While some patient-to-patient variation was observed, there was no clear difference in either the timing or intensity of the response between any of the outcome groups. Indeed, comparing the levels of each antibody at the first blood draw, there was no significant difference observed between ICU and non-ICU participants in this cohort (**Supplemental Figure 2C**).

While the antibody levels were highly correlated, there were some patient-specific differences, especially in the development of N-terminal versus C-terminal specific anti-Nucleocapsid antibodies. To better assess this, we performed principal component analysis (PCA) on the anti-Nucleocapsid antibody data, which identified 5 independent clusters (**Figure 4G, Supplemental Figure 2E**). Cluster 1, 2, and 5 corresponded to specimens with consistently low, medium, and high levels of anti-Nucleocapsid antibodies (C-terminal, N-terminal, and full-length, **Figure 4H**). Cluster 3, however, consisted of three participants with high antibody levels to the Nucleocapsid C-terminus, but low antibody levels to the Nucleocapsid N-terminus, while Cluster 4 consisted of three participants with high antibody levels to the N-terminus, but low antibody levels to the C-terminus (**Figure 4H**). Interestingly, levels of anti-Spike antibodies tracked more closely with the antibody levels against anti-Nucleocapsid C-terminus (higher in Cluster 3 and lower in Cluster 4, **Figure 4H**), though more specimens would be needed to verify this observation. Regardless, PCA cluster by serology did not significantly predict outcome (**Supplemental Figure 2F**), consistent with our prior analysis. Similarly, the presence of anti-HLA antibodies and plasma levels of PAI-1 were measured, but neither were significantly associated with outcome (**Supplemental Figure 2A, Supplemental Figure 2B**).

### Association of viral load with clinical measures and COVID-19 outcome

While early reports suggested associations between viral load and disease severity in COVID-19, subsequent reports have found little to no association [53]. Using a validated quantitative PCR (qPCR) assay for SARS-CoV-2 (N1 primer set, CDC assay [31]), we determined cycle threshold (Ct) values as a proxy for viral load in each of the nasopharyngeal swabs longitudinally collected from the study participants (**Figure 1A**). All study participants had detectable viral loads upon enrollment and generally showed a reduction in virus levels (increase in Ct values) over their hospital stay (**Figure 5A** and **Figure 5B**). The timing of this reduction was not always uniform, with several ICU participants and several participants who would ultimately die showing transient increases in viral load even 2 weeks after hospitalization (**Figure 5A**). While Ct values measured at or within 10 days of admission showed no significant difference by outcome (**Supplemental Figure S3A**), Ct values were significantly lower in non-ICU participants compared to ICU participants at or within 10 days of discharge (**Figure 5C**). Indeed, all non-ICU participants had detectable viral loads at or within 10 days of discharge while several ICU participants had at least one specimen with no detectable virus over the course of hospitalization (**Figure 5A** and **Figure 5B**).

**FIGURE 5.**
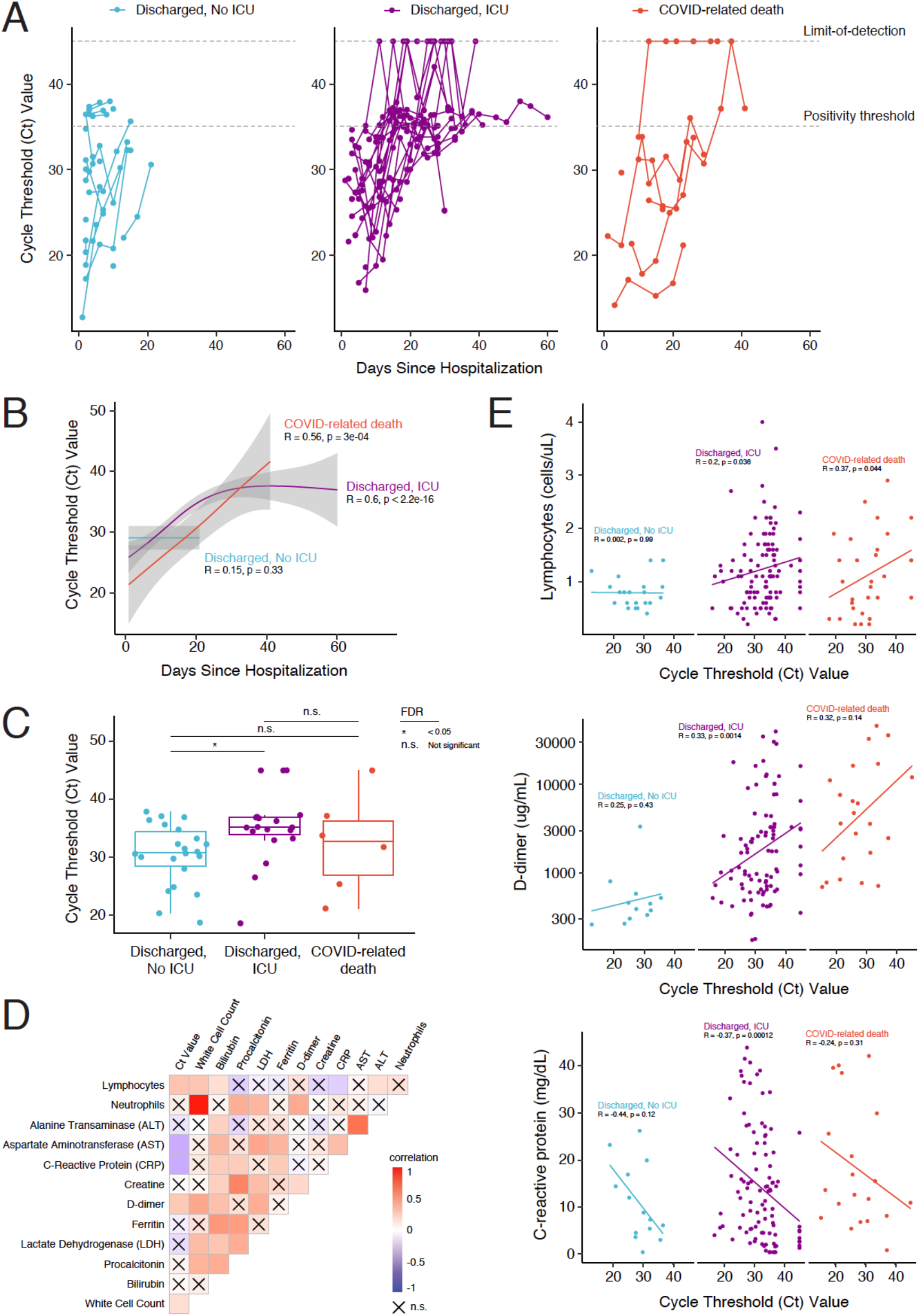
Analysis of correlations between viral load, clinical laboratory tests, and participant outcome. **A)** Plot of the quantitative PCR cycle threshold (Ct) values for SARS-CoV-2 N1 in nasopharyngeal swabs from each study participant relative to hospitalization time, separated by participant outcome. Specimens that did not amplify were assigned a value at the limit-of-detection. **B)** Generalized additive model fit to the average Ct value of hospitalized study participants over time separated by outcome. The correlation between average DI score and time during hospitalization is provided (R) alongside the p-value for each outcome group. **C)** Box plot of SARS-CoV-2 N1 Ct values from the final nasopharyngeal swab collected from each participant within 10 days of hospital discharge or death grouped by outcome (Wilcoxon rank sum test with Benjamini-Hochberg procedure to control FDR for multiple comparisons; n.s. = FDR >0.05, * = FDR <0.05). **D)** Heat map of Spearman correlation among longitudinally reported blood work variables over the entire course of hospitalization alongside Ct value. Value of the correlation coefficient is colored blue to red from more negatively to more positively correlated, respectively. Lack of significant correlation (p-value >0.05) is indicated by an X. **E)** Scatterplot of lymphocyte count (cells per microliter, top), D-dimer levels (microgram per milliliter, middle), and C-reactive protein levels (milligrams per deciliter, bottom) in blood versus time-matched Ct value. Study participants are separated by outcome group (no ICU care in blue, some ICU care in purple, COVID-related death in red) with the linear regression fit shown as a solid line of the same color (Spearman correlation coefficient and p-value shown).

Comparing Ct values in nasopharyngeal swabs with anti-Spike and anti-Nucleocapsid antibodies in time-matched patient plasma, there is a weak, but significant correlation with higher antibody titers correlated with higher Ct values. Antibodies targeting the Spike receptor-binding domain were the most highly correlated with Ct value (R = 0.5, p-value = 0.000065) while antibodies to the Nucleocapsid N-terminus were the least correlated (R = 0.28, p = 0.03) (**Supplemental Figure S3B** and **Supplemental Figure S3C**). Likewise, comparing Ct values with time-matched clinical laboratory test results identified several significant correlates (**Figure 5D**), including positive correlations with lymphocyte count, white cell count, and D-dimer levels, and negative correlations with aspartate aminotransferase (AST) and C-reactive protein (CRP) levels (**Figure 5E**). Notably, the factors most associated with disease severity (LDH level and neutrophil count, **Figure 4C**) were not significantly correlated with Ct values, while AST levels and lymphocyte counts were correlated with Ct values, but not disease severity.

### Analysis of viral genotype and intra-host diversity

As SARS-CoV-2 continues to diversify across the globe, several variations associated with increased viral load, viral transmission, and/or disease severity have been identified [54, 55]. To determine if viral genotype was significantly associated with outcome in this cohort, we attempted whole genome sequencing of viral isolates in all available nasopharyngeal swabs. Of the 238 swabs collected, 83 had adequate viral copies for sequencing (Ct value <30). Of these, 69 specimens from 34 independent participants yielded SARS-CoV-2 sequence of sufficient quality to assemble near complete genomes (at least 90% coverage, minimum read depth of 10 reads). Phylogenetic analysis revealed these viruses belonged to four primary clades, consistent with the population structure of the epidemic in Chicago during the study: Nextstrain clades 19A (2 sequences in 2 participants, representing Pango lineage A.3), 20A (13 sequences in 8 participants, representing Pango lineages B.1, B.1.378, and B.1.416), 20B (3 sequences in 1 participant, representing Pango lineage B.1.1.29), and 20C (51 sequences in 23 participants, representing Pango lineages B.1, B.1.162, B.1.521, B.1.313, B.1.422, and B.1.2) (**Figure 6A**). Longitudinal samples from the same participant clustered closely together, as expected, with 23 participants yielding identical consensus sequences at each timepoint. 9 participants, however, saw evidence of viral diversification with one or two mutations arising over time (**Figure 6A**). Clade membership was not significantly associated with any one outcome group (**Figure 6B**), nor did it correlate significantly with Ct value at the first sampled timepoint (**Figure 6C**).

**FIGURE 6.**
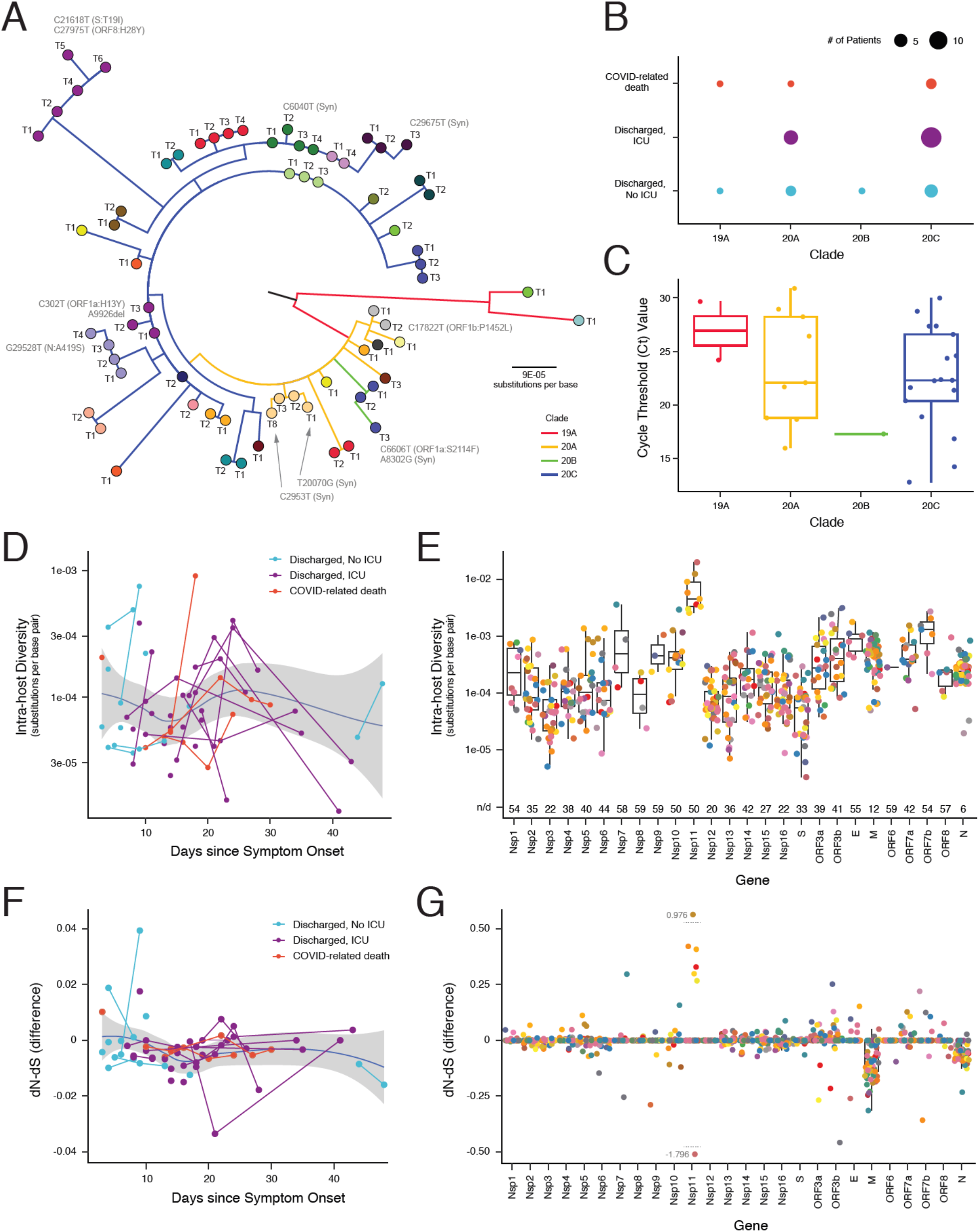
Analysis of correlations between viral genotype, intra-host diversity, and participant outcome. **A)** Phylogenetic tree of SARS-CoV-2 whole genome sequences from each study participant and timepoint. Each branch is colored by corresponding viral clade and each branch tip is colored by study participant; tips are labeled chronologically from timepoint 1 (T1) onward ordered clockwise. Changes in consensus viral sequence observed are labeled by nucleotide and corresponding amino acid change, when applicable. **B)** Dot plot depicting the clade membership of the virus isolated from each study participant by outcome group. **C)** Box plot comparing the SARS-CoV-2 N1 cycle threshold (Ct) value at timepoint 1 for each participant separated by clade. Tests for significance were performed with the Wilcoxon rank sum test with Benjamini-Hochberg correction for multiple comparisons (all comparisons not significant, p-value >0.05). **D)** Plot of the observed intra-host diversity (substitutions per base pair) across the entire SARS-CoV-2 genome from each study participant relative to days since symptom onset and colored by participant outcome. Samples from the same participant over time are linked by lines. A loess curve fit to the average intra-host diversity observed over time is overlaid with error. **E)** Box plot of the intra-host diversity observed within each indicated open reading frame in each participant specimen. Dots are colored by study participant. The number of specimens in which no intra-host diversity was observed is indicated on the bottom (n/d). **F)** Plot of the difference between the number of nonsynonymous substitutions per non-synonymous site (dN) and the number of synonymous substitutions per synonymous site (dS) observed across the SARS-CoV-2 genome from each study participant relative to days since symptom onset and colored by participant outcome. Samples from the same participant over time are linked by lines. A loess curve fit to the average dN-dS observed over time is overlaid with error. **G)** Box plot of the dN-dS observed within each indicated open reading frame in each participant specimen. Dots are colored by study participant.

Given the appearance of mutations in the consensus sequence of some patient isolates over time, we investigated the degree of intra-host diversity in each participant at each timepoint. While the amount of intra-host diversity varied isolate to isolate, there was no clear trend either as a function of participant outcome or as a function of time since symptom onset (**Figure 6D**). On the contrary, the viral population diversity of any given isolate was roughly 0.0001 substitutions per base pair. Diversity did vary by open-reading frame (ORF), with a majority of sequences showing some diversity in nucleoprotein (N, n = 63), membrane (M, n = 57), and nsp12 (n = 49) sequences (**Figure 6E**). While nsp11 and ORF7b showed the highest number of substitutions per base pair, this was in fewer isolates (n = 9 and n = 7, respectively) and was largely driven by small ORF size.

To determine if this diversity reflected any significant negative selection or positive selection, we calculated the difference in the number of nonsynonymous substitutions per nonsynonymous site (dN) and the number of synonymous substitutions per synonymous site (dS) (dN-dS). A bias in dN-dS towards synonymous mutations (negative value) is suggestive of negative selective pressure, while a bias in dN-dS towards nonsynonymous mutations (positive value) is suggestive of positive selective pressure [56, 57]. Overall, we find that dN-dS across the entire genome does not change notably by outcome or as a function of time since symptom onset (**Figure 6F**). dN-dS does vary as a function of ORF with the two most frequently changed ORFs (M and N), showing a significant bias towards synonymous mutations, suggestive of negative selective pressure operating at the intra-host level (**Figure 6G**). On the other hand, Nsp11 and the immune regulatory factors ORF3a/b and OR7a/b showed some bias towards a positive dN-dS, suggestive of positive selection, though this was not apparent in all participants (**Figure 6G**).

### Model of COVID-19 Outcome

A multinomial logistic model with the three COVID-19 outcome groups as the outcome variable was constructed to assess the relative predictive value of each measured parameter at or near (within 10 days) the time of hospital admission. Based on the univariant analyses above, candidate predictors with p-value less than 0.1 were identified for multivariable assessment including: time since symptom onset, sex, age, BMI, LDH levels, lymphocyte count, CRP levels, neutrophil count, white blood cell count, DI score, and N1 Ct value. SpO_2_ and FiO_2_ were excluded from these analyses as these parameters were used in part by clinical staff to dictate ICU admission during the course of the study. Only study participants with complete datasets for all measurements were included in the model (n = 34).

Of all examined parameters, DI score at admission was by far the most significant predictor (p-value <0.0001), yielding an accuracy rate of 0.88, even when considered independently from all other variables. Given that DI score is proprietary to the Epic medical record systems and only available in hospital settings that use that software, we reran the model excluding DI score. After fitting the initial model (**Figure 7A**), we performed a stepwise selection using both forward inclusion and backward elimination of the candidate predictors. After selection, only BMI, lymphocyte count, and neutrophil count were maintained in the final model, which had an accuracy rate of 0.82 in predicting all outcome groups (**Figure 7A**). A detailed analysis performed on the outcome predictor effect for each variable indicated that BMI and lymphocyte count were the primary drivers differentiating between ICU and non-ICU individuals, while neutrophil count enabled discrimination of COVID-related deaths (**Figure 7B**). Although the predictive capability of this model is limited due to small sample size, this model was able to correctly predict all COVID-19 related deaths included in the model dataset. Furthermore, it performed significantly better than a model using only the significantly associated demographic variables (age, sex, and BMI), which yielded an accuracy rate of only 0.62.

**FIGURE 7.**
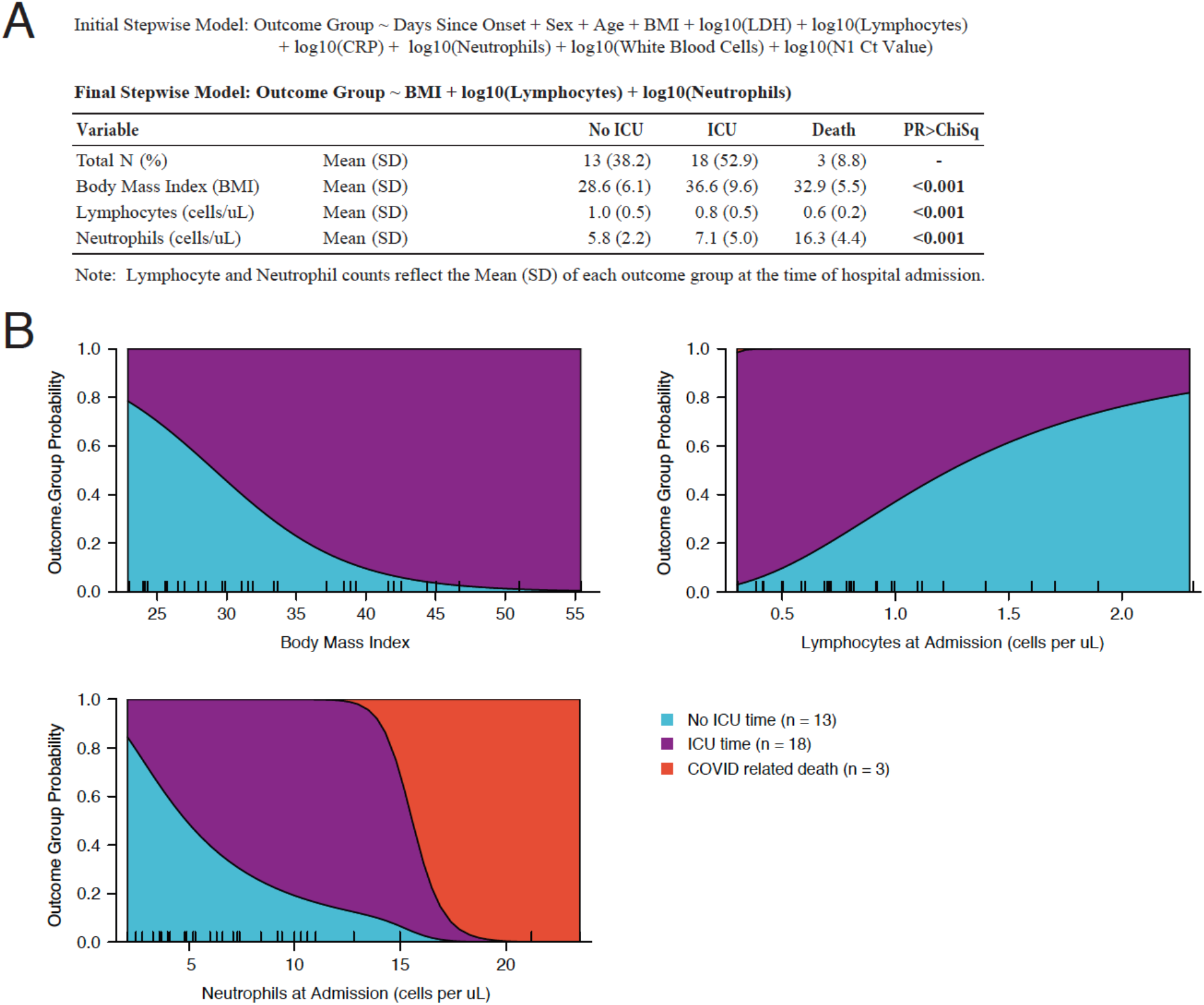
Stepwise Regression Model of Participant Outcome by Admission Data. **A**) Initial and final stepwise model after Akaike information criterion (AIC) model selection. The distribution and statistical significance of study participant body mass index (BMI), blood lymphocyte count on admission, and blood neutrophil count on admission towards predicting outcome in the final model is depicted in the table. All continuous variables are represented as mean (standard deviation). **B)** Probability of each outcome determined by BMI, lymphocyte count at admission, and neutrophil count at admission in the final model. The BMI and cell counts of each included study participant are indicated by a tick mark along the x-axis.

## DISCUSSION

*De novo* prediction of a patient’s risk of severe COVID-19 and poor outcome remains imprecise, despite the identification of several independently associated factors, including: demographic factors, such as age and sex [1], clinical factors, such as BMI, known co-morbidities, and respiratory rate [9, 58], virological factors, such as viral load and viral genotype [53]; and serological factors, such as inflammatory markers, antibody responses, and autoimmune complications [59, 60]. A number of composite measures of patient deterioration have likewise been developed to assist with clinical management of disease, but no one measure is utilized consistently across disparate health systems [16]. Furthermore, few studies have directly compared and validated independent associations in the context of a single cohort. Here, we have assessed and compared a series of demographic, clinical, virological, and serological parameters for association with patient outcome in a single cohort of hospitalized COVID-19 participants in Chicago, Illinois during the first wave of the pandemic in early 2020.

A total of 58 participants were longitudinally sampled and compared between three hospital outcome groups: discharged without ICU care, discharged after some ICU care, and death due to COVID-19 related complications. As expected, and as reported by other studies, higher age, male sex, and higher body mass index (BMI) were associated with ICU admission. Neither race nor ethnicity were significantly associated with hospital outcomes in this cohort, though we would note that our study was limited to individuals admitted within a single system, negating some of the potential impacts due to disparities in access and quality of health care. We furthermore saw no significant impact of co-morbidities on outcome among already hospitalized participants, though the average number of co-morbidities was relatively high (roughly 3) in all outcome groups. Although this study did not include a non-hospitalized control group for comparison, it is possible that the number of co-morbidities may be a better predictor of requirement for hospitalization than outcome after hospitalization.

Several longitudinally assessed measurements taken during routine clinical care were also found to be significantly associated with outcomes. As previously reported, neutrophil and white blood cell counts over the course of hospitalization were associated with the need for ICU care [50, 61, 62]. Both longitudinally assessed composite measures of deterioration, MEWS and DI score, likewise reached significantly higher maximum and median levels in participants who spent time in the ICU. DI score, in particular, was starkly different between ICU and non-ICU participants, with the median DI score showing significant differences even between individuals in the ICU who would recover versus die. Modeling of these two groups showed almost identical behavior of the DI score over the first 10 days of hospitalization, after which time individuals who would eventually recover displayed a gradual decrease while those who died showed a gradual increase. This was one of the only measures in this study to show significant differences for death as an outcome, suggesting that tracking of not just daily, but cumulative measures of these deterioration indices may be useful for risk stratification.

While these factors all correlated over the course of hospitalization, fewer measures were found to be predictive of outcome upon hospital admission. Of all the clinical laboratory tests examined, only LDH levels showed a significant association with ICU admission. LDH is a commonly used marker of tissue damage, suggesting these participants may have been experiencing more severe disease even at the time of hospitalization. Importantly, this is not a function of symptom duration prior to hospital presentation as there were no significant differences between outcome groups in the time from symptom onset to the time of hospital admission. The 4C mortality score, which was originally developed to predict patient outcome at admission, likewise showed significant differences between non-ICU and ICU participants at hospital admission, though it was not significantly different between ICU participants who recovered and those who died. No parameter monitored at hospital admission was significantly associated with death in this cohort, emphasizing the importance of longitudinal monitoring and further study.

Viral load was not associated with outcome group in this study, consistent with prior reports [63]. Indeed, non-ICU participants had significantly higher viral loads at the time of discharge compared to ICU participants, though this may simply be due to shorter hospital stays. Likewise, the significant correlates of outcome (LDH and neutrophil count) were not significantly associated with viral load. Taken together, these results suggest that inflammatory responses in response to infection and markers of lung damage are more important than baseline viral load to predict outcomes. Interestingly, viral load did correlate with other measured parameters in this study, including positively with AST levels and negatively with total lymphocyte count and antibody titer, none of which were associated with outcome. Examination of these competing markers in a larger cohort would be warranted to determine how such commonly performed lab tests could be repurposed to not only inform disease severity predictions, but also risk of transmission. In addition, these markers may be useful to look at in populations that received antiviral and/or anti-inflammatory therapy for COVID-19 to define predictive biomarkers for selecting optimal therapy for individual patients.

Most patients infected with SARS-CoV-2 elicit robust antibody responses to both the S and N proteins [64–66], though the significance of timing and strength of those responses to predicting clinical outcome remains unclear [52, 53, 67]. Here, we likewise observed that a majority of participants developed antibodies to both the N and S protein, usually within two weeks of symptom onset. These responses were not observed to differ by outcome group, though they were broadly correlated with viral load over time. We furthermore observed unique clusters of participants with differential antibody responses by antigen, most notably in response to different domains of the nucleocapsid protein. Previous reports have suggested that C-terminal fragments of N are highly immunogenic in more severely ill hospitalized patients as compared to non-hospitalized controls [64, 68]. While this was not observed here, the study was limited to only hospitalized individuals. Interestingly, antibody responses to the Spike RBD were more closely correlated with responses to the C-terminal domain of N, though this would need to be validated in larger cohort studies.

Viral genotype in the cohort was broadly reflective of the epidemiological trends in the city of Chicago at the time of sample collection and showed no significant correlation with outcome group [69]. No significant associations between SARS-CoV-2 clade and viral load were detected either, though only two of the isolates were from clade 19A. These two isolates were the only ones to have Spike D614 as opposed to the D614G mutation, which had been previously shown to influence viral loads in patient upper airways [54, 55]. Both of these samples did have lower overall viral loads, but a larger cohort would be needed to assess significance. Sample collection was completed prior to the emergence of the now prevalent variants-of-concern, several of which have been associated with elevated viral loads, increased transmission, and potentially worse patient outcomes [18–25]. Overall viral genetic diversity in the population was lower earlier in the epidemic while this study was ongoing compared to diversity observed today among currently circulating variants. While this limited potential linkage of viral variation with phenotype here, it was also beneficial in removing one potential confounder when assessing correlation with other variables. Going forward, this cohort may prove valuable as an outgroup for more recent and future patient cohorts to determine the impact of emerging variants on disease severity and patient outcomes.

Longitudinal monitoring of viral genotype revealed nine instances of intra-host evolution and the emergence of new, predominant mutations in the viral population. Most of these changes were rare mutations that are not detected globally at the consensus level indicating random or host-specific adaptation events. However, one participant developed a mutation in the position 19 of the Spike protein (T19I) that, although uncommon (<0.5% of global sequences), is in the same position as the mutation T19R in the recently emerging Clade 21A. These mutations all occurred at later timepoints, though it is unclear if these were driven by humoral responses or arose due to random chance in the setting of ongoing viral replication in the host. Overall viral genetic diversity remained relatively constant throughout the course of hospitalization, even up to 45 days after admission, and did not vary significantly by outcome. Select ORFs, such as M and N, did show evidence of negative selection pressure as measured by dN-dS, though most ORFs showed little selection bias. The low variability observed, and the predominance of synonymous mutations within this variability, may be indicative of low intra-host selective pressure exerted by the immune system during the course of acute infection. Comparing the changes in viral genotype that arose in these participants with host-specific humoral and innate responses will be important to better understand the factors that drive evolution of SARS-CoV-2, particularly in cases of prolonged hospitalization. This emphasizes the need for effective antivirals to suppress viral replication to avoid the evolution of immune evasive variants. This is particularly important in cases of prolonged replication as seen in immunocompromised patients [70].

Comparing outcome correlates head-to-head as predictors of disease outcome at hospital admission, the DI score was found to be the most predictive measure, accurately calling 88% of outcomes in a logistic model. Retrospective analysis of this measure as a predictor of COVID-19 outcome in hospital systems that use this software needs to be performed across a larger and more diverse sample size to assess its value in risk management, though this is likely to be complicated by hospital systems that use this index in part to determine level of care. Unfortunately, the proprietary nature of the score prevents further dissection of the factors driving predictability. That being said, a multinomial logistic model including BMI, lymphocyte count at admission, and neutrophil count at admission was nearly as accurate, calling 82% of outcomes correctly in this cohort. Given that these values are more universally available across hospital systems, validation of this model in larger cohorts may lead to more broadly applicable risk stratification strategies. Notably, both models outperformed the demographics only model, which had an accuracy rate of 62%.

Despite its strengths, this study has several limitations. This is a single center study and as such our population may be different from the general population. That said, during this phase of the pandemic, we were a tertiary referral hospital that continued to accept patients from both our usual catchment area but also referrals in from other hospitals around the Chicagoland area. The study included the first wave of the pandemic and as such there was significant variability in the therapies that were given to the participants. That said, few of the participants received therapies that were subsequently found to be clinically beneficial. As such, the experience reflects the natural history of illness with general supportive measures. Nevertheless, continued exploration of the trends observed here in larger, multi-institutional cohorts is required. Lastly, the study did not include a non-hospitalized control group for comparison and utility of the identified markers to predict hospitalization or other outcomes cannot be assessed.

In sum, this study found that the 4C mortality score and LDH levels at the time of admission were predictive of admission to the ICU and should be examined in larger cohorts for use in clinical risk management. Validation of a novel score based on BMI, lymphocyte count, and neutrophil count on admission may yield a useful tool for predicting outcomes of hospitalized patients. While not assessed in this study, the role of these factors and others to predict optimal therapy selection are also needed. Continued exploration of these trends in larger cohort data will be essential to translate these findings into improved strategies for clinical care and risk stratification.

## Data Availability

All data will be available as supplemental material; sequencing data is available on GSAID.

## ACKNOWLEDGEMENTS

This research was supported in part through the computational resources and staff contributions provided for the Quest high performance computing facility at Northwestern University, which is jointly supported by the Office of the Provost, the Office for Research, and Northwestern University Information Technology. Clinical data collection was supported in part by the Northwestern Medicine Enterprise Data Warehouse. Sample collection was supported by a COVID-19 pilot grant from the Northwestern University Clinical and Translational Science Institute (NUCATS, NIH grant UL1 TR001422). Additional funding for this work was provided by: a Northwestern Institute for Global Health Catalyzer Research Fund award (R.L.R.); a NUCATS COVID-19 Collaborative Innovation Award (R.L.R); a Dixon Translational Research Grant made possible by the generous support of the Dixon Family Foundation (E.A.O., J.F.H.); a CTSA supplement to NCATS UL1 TR002389 (J.F.H., E.A.O., R.L.R.); a supplement to the Northwestern University Cancer Center P30 CA060553 (J.F.H., K.E.R.B., K.J.F.S.); the Center for Structural Genomics of Infectious Diseases at Northwestern University (NIH/NIAID Contract # HHSN272201700060C, K.J.F.S.), the Gilead Sciences Research Scholars Program in HIV (J.F.H.); the NIH-supported Third Coast CFAR P30 AI117943 (R.L.R., J.F.H.); NIH grant U19 AI135964 (E.A.O.); NIH grant 3UL1 TR001422-06S4 (E.M.M. and A.R.D.); through a generous contribution from the Walder Foundation (J.F.H., E.A.O., R.L.R.); and through a generous gift from Dr. Andrew Senyei and Noni Senyei (E.M.M. and A.R.D.). The funding sources had no role in the study design, data collection, analysis, interpretation, or writing of the report.

## AUTHOR CONTRIBUTIONS

Conceptualization: L.M.S., R.L.R., K.J.F.S., E.A.O., M.G.I., J.F.H.; Sample Acquisition: L.M.S., M.G., S.L.K., M.G.I., J.F.H.; Data Collection: L.M.S., R.L.R., M.G., S.L.K., J.P.V., N.L.R., M.E., E.L., K.E.R.B., A.R.D., C.J.A., E.A.O., M.G.I., J.F.H.; Formal analysis: L.M.S., R.L.R., E.A.O. J.F.H.; Supervision: R.L.R., E.M.M., A.R.T., D.E.V., K.E.R.B., A.R.D., K.J.F.S., C.J.A., E.A.O., M.G.I., J.F.H.; Funding acquisition: R.L.R., E.M.M., A.R.T., D.E.V., K.E.R.B., A.R.D., K.J.F.S., C.J.A., E.A.O., M.G.I., J.F.H.; Writing: R.L.R., K.E.R.B., K.J.F.S., E.A.O., M.G.I., J.F.H. Editing: R.L.R., K.E.R.B., K.J.F.S., E.A.O., M.G.I., J.F.H.

---

**SUPPLEMENTAL FIGURE 1.**
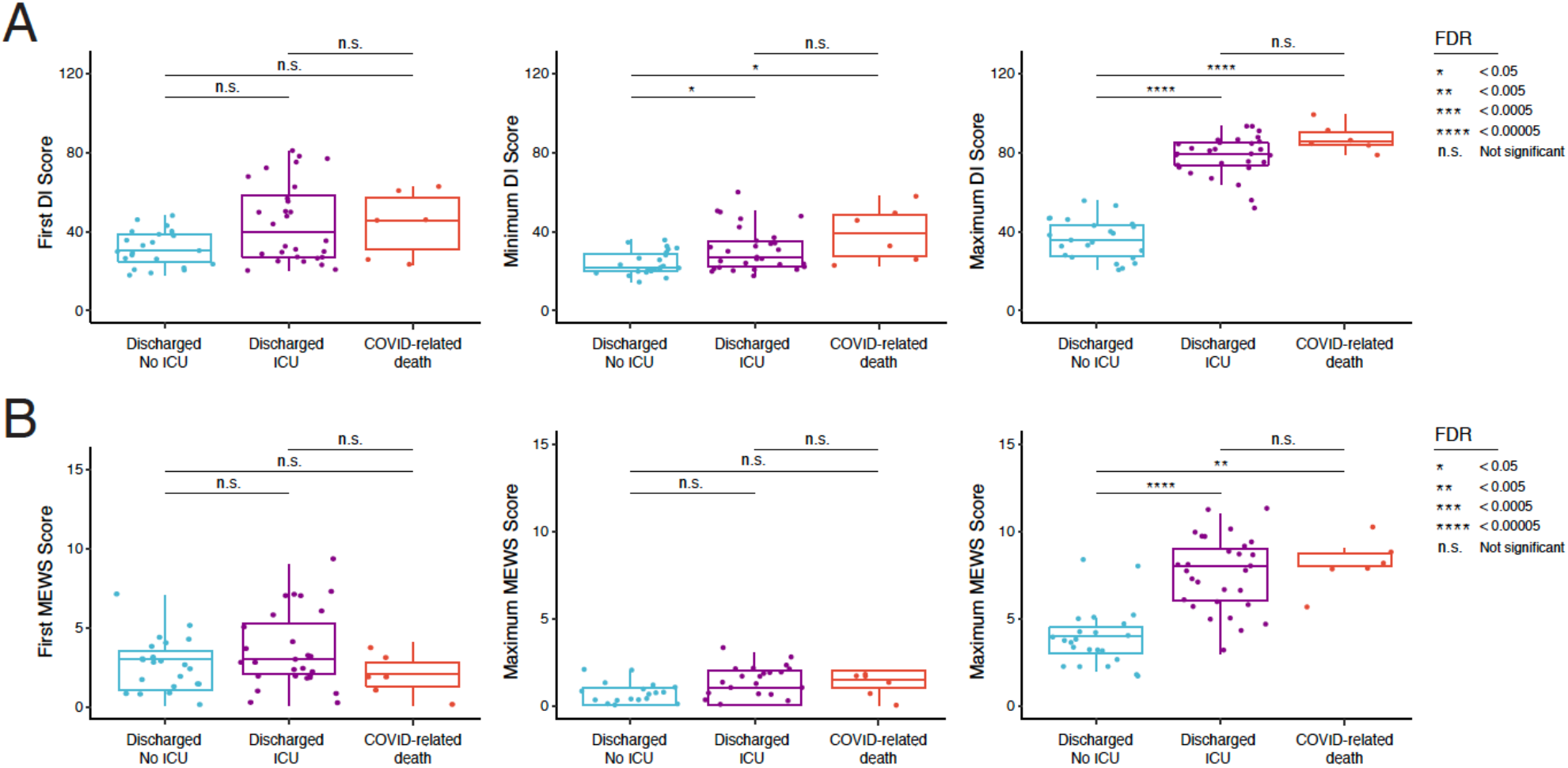
Analysis of correlations between DI Score and MEWS with participant outcome at different times during hospitalization. Box plots comparing the **A)** Deterioration Index (DI) scores and **B)** Modified Early Warning Scores (MEWS) by participant outcome group including the first reported score (left), the maximal reported score (center), and the minimal reported score for each study participant (right). Significance between groups in all box plots was tested by Wilcoxon rank sum test with Benjamini-Hochberg procedure to control FDR for multiple comparisons; n.s. = FDR >0.05, * = FDR <0.05, ** = FDR <0.005, *** = FDR <0.0005, **** = FDR <0.00005.

**SUPPLEMENTAL FIGURE 2.**
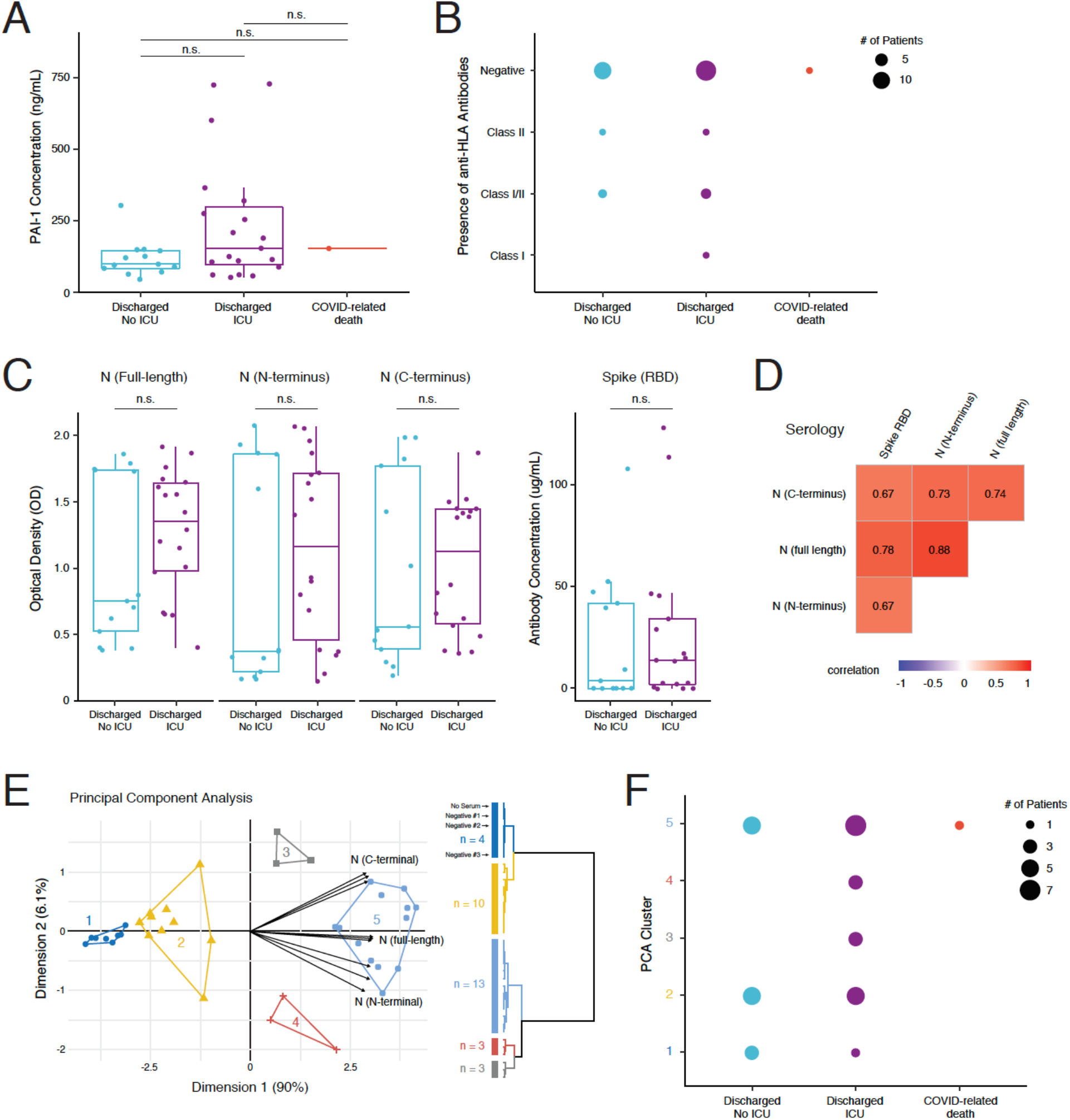
Analysis of correlations between PAI-1 levels, anti-HLA antibody levels, serology, and participant outcome. **A)** Box plot of PAI-1 concentration (nanograms per milliliter) in the plasma of study participants at their first draw by outcome group (Wilcoxon rank sum test with Benjamini-Hochberg procedure to control FDR for multiple comparisons, n.s. = FDR >0.05). **B)** Dot plot depicting the presence or absence of antibodies against HLA Class I or Class II in the serum of study participants at their first draw by outcome group. **C)** Box plots comparing the OD value of antibody in patient plasma recognizing the full-length (left), N-terminus (center), or C-terminus (right) of the Nucleocapsid (N) as determined by ELISA by outcome group at the time of the first blood draw. The box plot on the far right depicts the concentration of IgG antibody recognizing Spike RBD in each specimen by outcome group at the time of the first blood draw. Tests for significance were performed with the Wilcoxon rank sum test with Benjamini-Hochberg procedure to control FDR for multiple comparisons (n.s. = FDR >0.05). **D)** Heat map of Spearman correlation among antibodies recognizing Spike RBD, the C-terminus of N, the N-terminus of N, and full-length N. The value of the correlation coefficient is colored blue to red from more negatively to more positively correlated, respectively. **E)** Principal component analysis (PCA) plot depicting study participant grouping by N antibody levels, including vector contributions of each replicate of each variable. The clustering of each study participant obtained by agglomerative hierarchical clustering on the PCA results is depicted on the right. **F)** Dot plot depicting the PCA cluster of each study participant by outcome group.

**SUPPLEMENTAL FIGURE 3.**
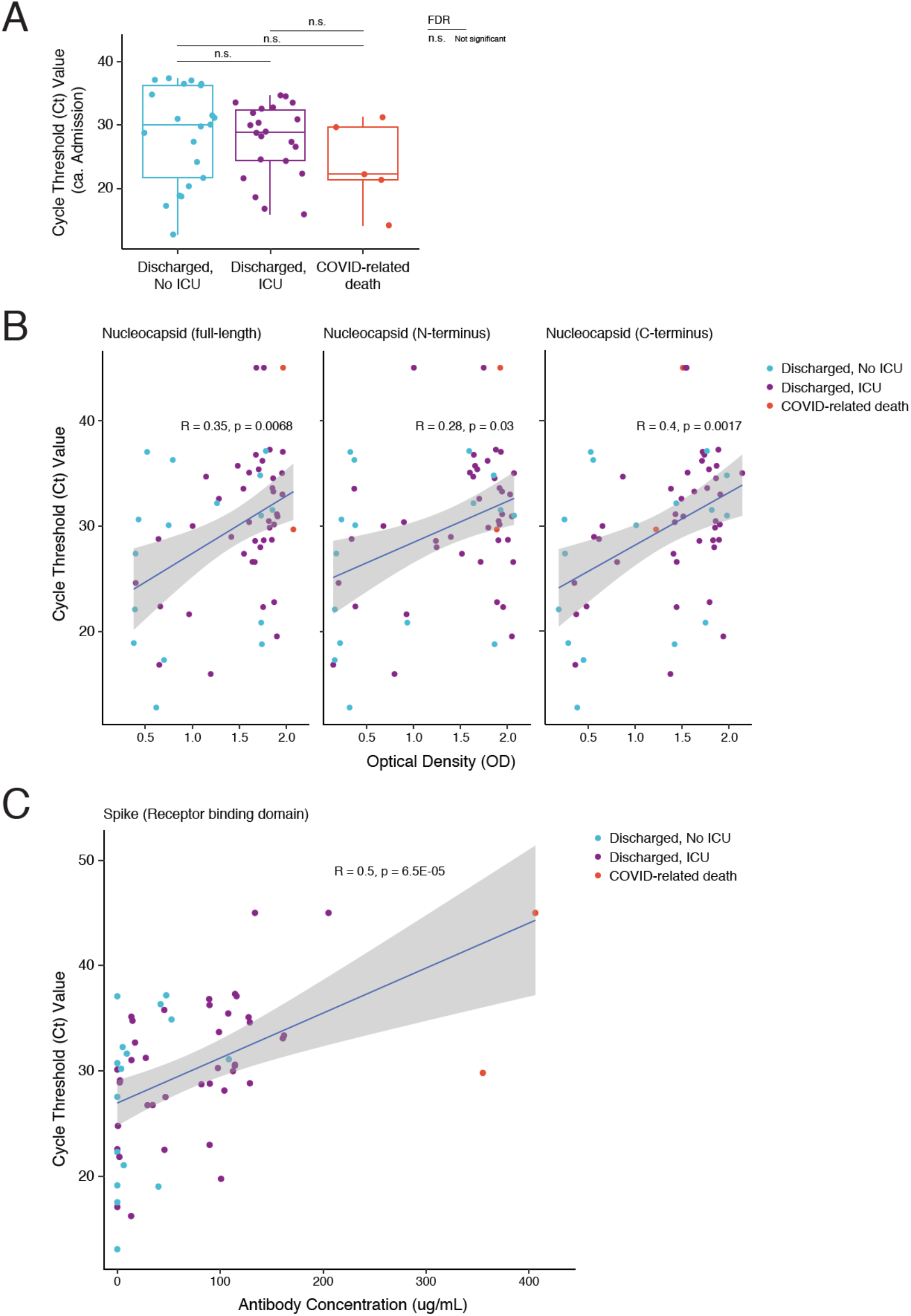
Analysis of correlations between viral load and serology data. **A)** Box plot of SARS-CoV-2 N1 cycle threshold (Ct) values from the first nasopharyngeal swab collected within 10 days of hospital admission for each participant grouped by outcome (Wilcoxon rank sum test with Benjamini-Hochberg procedure to control FDR for multiple comparisons; n.s. = FDR >0.05). **B)** Scatter plot of the Optical Density (OD) value reflective of the concentration of antibody recognizing the full-length (left), N-terminus (center), or C-terminus (right) of the Nucleocapsid (N) protein in the blood as determined by ELISA versus time-matched SARS-CoV-2 N1 Ct values from paired nasopharyngeal swabs. Study participants are colored by outcome group (no ICU care in blue, some ICU care in purple, COVID-related death in red) with the linear regression fit and error overlaid (Spearman correlation coefficient and p-value shown). **C)** Scatterplot of the concentration of IgG antibody (micrograms per milliliter) recognizing the receptor binding domain (RBD) of Spike as determined by ELISA versus time-matched SARS-CoV-2 N1 Ct values from paired nasopharyngeal swabs. Study participants are colored by outcome group as above with the linear regression fit and error overlaid (Spearman correlation coefficient and p-value shown).

